# Broad anti-SARS-CoV-2 antibody immunity induced by heterologous ChAdOx1/mRNA-1273 prime-boost vaccination

**DOI:** 10.1101/2021.12.13.21267598

**Authors:** Chengzi I. Kaku, Elizabeth R. Champney, Johan Normark, Carl E. Johnson, Clas Ahlm, Mrunal Sakharkar, Margaret E. Ackerman, Mattias N. E. Forsell, Laura M. Walker

## Abstract

Heterologous prime-boost immunization strategies have the potential to augment COVID-19 vaccine efficacy and address ongoing vaccine supply challenges. Here, we longitudinally profiled SARS-CoV-2 spike (S)-specific serological and memory B cell (MBC) responses in individuals receiving either homologous (ChAdOx1:ChAdOx1) or heterologous (ChAdOx1:mRNA-1273) prime-boost vaccination. Heterologous mRNA booster immunization induced significantly higher serum neutralizing antibody and MBC responses compared to homologous ChAdOx1 boosting. Specificity mapping of circulating S-specific B cells revealed that mRNA-1273 booster immunization dramatically immunofocused ChAdOx1-primed responses onto epitopes expressed on prefusion-stabilized S. Monoclonal antibodies isolated from mRNA-1273-boosted participants displayed higher binding affinities and increased breadth of reactivity against variants of concern (VOCs) relative to those isolated from ChAdOx1-boosted participants. Overall, the results provide fundamental insights into the B cell response induced by ChAdOx1 and a molecular basis for the enhanced immunogenicity observed following heterologous mRNA booster vaccination.

---

Multiple safe and effective COVID-19 vaccines have been developed at an unprecedented scale and speed. However, waning vaccine-induced immunity and the emergence of SARS-CoV-2 VOCs with increased neutralization resistance have limited the effectiveness of currently available vaccines (*1-3*). Furthermore, on-going vaccine supply shortages and interruptions have slowed the pace of global mass-vaccination efforts. One significant interruption in vaccine distribution was driven by evidence of rare but serious thrombotic events associated with ChAdOx1 nCoV-19/AZD1222 (hereafter referred to as ChAdOx1) immunization, which led to a halt in ChAdOx1 booster vaccinations in certain age groups (*4, 5*). To fully immunize individuals who received a single dose of ChAdOx1 but were not eligible for a ChAdOx1 boost, several European governments recommended a switch from the clinical trial validated homologous ChAdOx1 booster to heterologous booster vaccination with mRNA-1273 (Moderna) or BNT162b2 (Pfizer/BioNTech). Recent studies have shown that heterologous ChAdOx1/mRNA prime-boost immunization induces higher serum neutralizing antibody titers and confers increased levels of protection relative to homologous ChAdOx1 dosing, but the molecular basis for this difference in immunogenicity remains unknown (*6-10*).

All COVID-19 vaccines in late-stage clinical trials, approved, or authorized for emergency use are based on the SARS-CoV-2 spike protein, which plays a key role in viral entry and is the only known target for neutralizing antibodies. The S protein exists on the surface of the virion in a metastable prefusion conformation, and binding of the receptor binding domain (RBD) to the host cell receptor, angiotensin converting enzyme 2 (ACE2), triggers shedding of the S1 subunit and transition of the S2 subunit to a highly stable postfusion conformation (*11*). Early structural and biochemical studies demonstrated that a stabilized version of the spike ectodomain that contains two consecutive proline substitutions in the S2 subunit (S-2P) prevents the transition from the prefusion to postfusion state and leads to higher expression levels in vitro and enhanced immunogenicity in animal models (*12, 13*). Several COVID-19 vaccines encode S-2P, such that the protein maintains the prefusion conformation and avoids S1 shedding (e.g., mRNA-1273, BNT162b2, and Ad26.COV2.S), whereas others express full-length wild-type (WT) S (e.g. ChAdOx1, Sputnik V, and CoronaVac), which likely leads to expression of both pre- and post-fusion conformations of S. In this study, we comprehensively interrogated serological and circulating B cell responses induced by prime immunization with an adenovirus vector-based vaccine encoding WT S (ChAdOx1) and following either a second dose of ChAdOx1 or an mRNA vaccine expressing S-2P (mRNA-1273).

We recruited 55 healthcare workers at the University Hospital of Northern Sweden receiving either homologous ChAdOx1:ChAdOx1 or heterologous ChAdOx1:mRNA-1273 prime-boost vaccination for blood donation (Table S1). The volunteers ranged from 28 to 62 years old, with median ages of 46.5 and 42 for ChAdOx1:ChAdOx1 and ChAdOx1:mRNA-1273 cohorts, respectively. Twenty-seven percent (15/55) of participants were men and 73% (40/55) were women. None of the volunteers had a history of prior infection with SARS-CoV-2. Study participants received one dose of ChAdOx1 and 9-12 weeks later received a second dose of either ChAdOx1 (n=28) or mRNA-1273 (n=27). We collected the first blood sample on the day of booster immunization to analyze ChAdOx1-primed immune responses and a second blood sample 7-10 days following the second dose to study the early secondary B cell response induced by homologous or heterologous booster vaccination (Fig. 1A).

**Figure 1.**
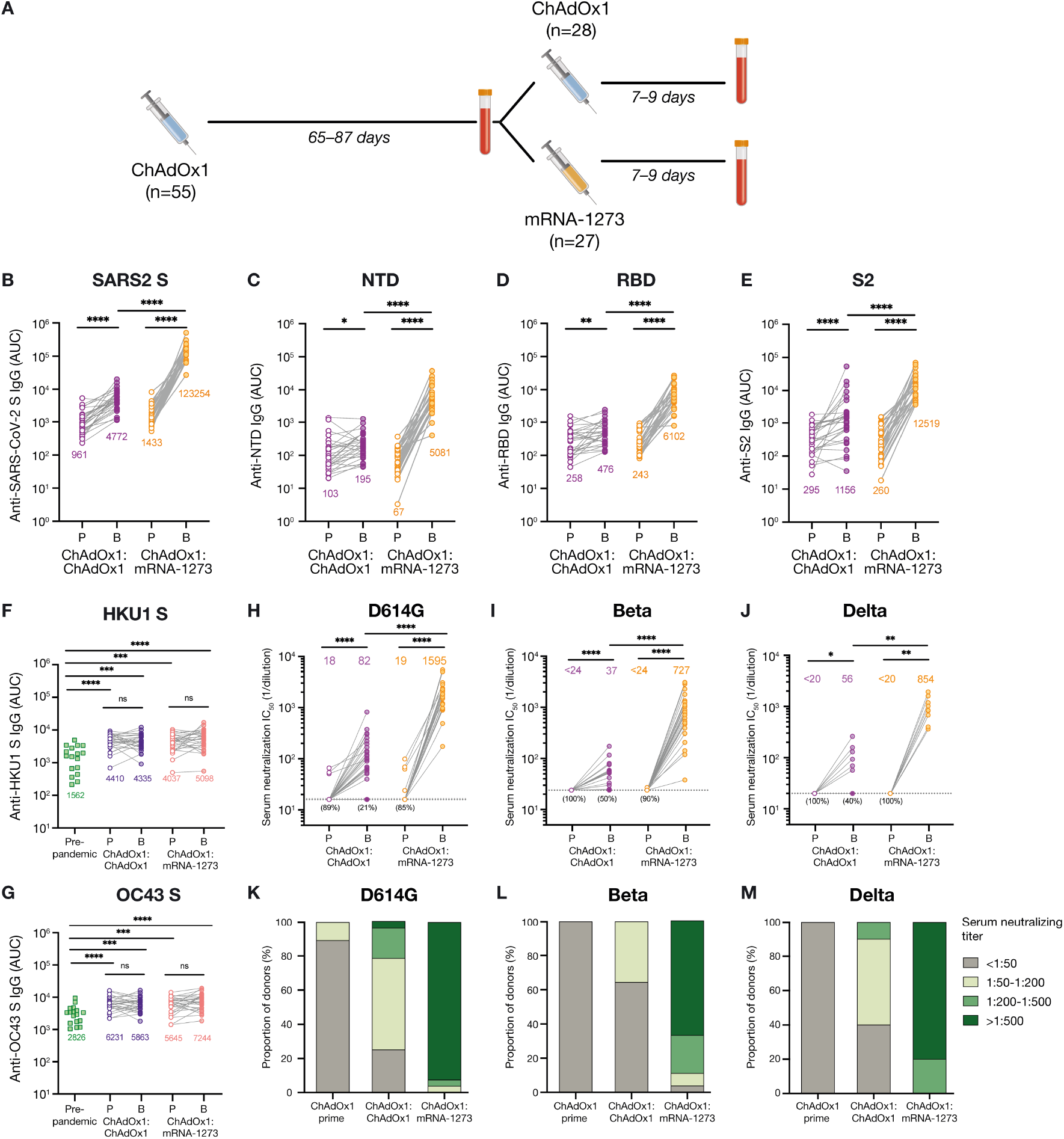
Serum binding and neutralizing activity following homologous and heterologous prime-boost vaccination. **(A)** Immunization and blood draw schedule. **(B-G)** Serum IgG binding to recombinant SARS-CoV-2 S (B), NTD (C), RBD (D), prefusion-stabilized S2 (E), HKU1 S (F), and OC43 S (G), as assessed by ELISA. Binding of pre-pandemic donor sera (N=17) is shown for comparison in F-G. Geometric mean AUC values are shown below data points. **(H-J)** Serum neutralizing activity against authentic SARS-CoV-2 D614G (H) and Beta/B.1.351 (I) measured by plaque reduction assay (N=27-28 donors from each cohort), and Delta/B.1.617.2 (J) measured by cytopathic effect (CPE)-based colorimetric microneutralization assay (N=10 donors from each cohort). Values below the dotted line indicate the percentage of samples with serum neutralizing titers below the limit of detection. **(K-M)** Proportion of donors with serum neutralization titers within the indicated stratifications for D614G (K), Beta (L), and Delta (M). Statistical comparisons between prime and boost were determined by Wilcoxon pair-matched rank sum test. Statistical comparisons across groups were determined by two-tailed Mann Whitney U test with Bonferroni correction (B-E and H) or two-sided Kruskal Wallis test by ranks with subsequent Dunn’s multiple comparisons (F-G). \**P* < 0.05, \*\**P* < 0.01, \*\*\**P* < 0.001, ****P<0.0001. P, Prime; B, Boost; AUC; area under the curve; LOD, limit of detection; ns, non-significant.

We first evaluated serum IgG binding activity in the samples collected at both pre- and post-boost timepoints. All participants mounted weak but detectable SARS-CoV-2 S-specific serum IgG binding responses following the first dose of ChAdOx1, and homologous booster vaccination resulted in a small but significant (4.6-fold) increase in serum IgG binding antibodies (Fig. 1B and Fig. S1). In contrast, heterologous booster vaccination with mRNA-1273 led to a dramatic (86-fold) increase in S-specific serum IgG binding responses (Fig. 1B and Fig. S1). Consistent with the responses observed for full-length S, ChAdOx1 prime immunization elicited weak serum IgG binding activity to recombinant receptor binding domain (RBD), N-terminal domain (NTD), and prefusion-stabilized S2 subdomains (geometric mean AUC ranging 67-321), and homologous and heterologous booster immunization enhanced these responses by 2-4-fold and 25-77-fold, respectively (Fig. 1C-E).

Previous studies have shown that cross-reactive MBCs induced by endemic β-CoVs are activated and expanded following primary SARS-CoV-2 infection and mRNA vaccination (*14, 15*). To determine whether homologous ChAdOx1:ChAdOx1 and/or heterologous ChAdOx1:mRNA-1273 prime-boost immunization regimens elicited seasonal β-CoV cross-reactive antibody responses, we assessed serum IgG binding activity to recombinant OC43 and HKU1 S proteins at both sampling timepoints. At the pre-boost time point, we detected significantly elevated serum antibody titers to both OC43 and HKU1 relative to pre-pandemic sera samples, suggesting recall of pre-existing cross-reactive MBCs induced by seasonal β-CoV infections (Fig. 1F, G). However, this response was not further amplified following homologous or heterologous booster immunization. Thus, it appears that ChAdOx1 prime immunization induces an original antigenic sin-like “back-boost” of cross-reactive antibodies to seasonal β-CoVs in SARS-CoV-2 naïve individuals.

We next evaluated serum neutralizing activity against authentic SARS-CoV-2 D614G, B.1.617.2/Delta, and B.1.351/Beta at both pre- and post-boost timepoints. We selected Delta and Beta VOCs due their prevalence and reduced susceptibility to neutralization by convalescent and vaccinee sera (*16, 17*). At 3 months post ChAdOx1 prime immunization, the vast majority of donors (87%) displayed weak or undetectable serum neutralizing activity against SARS-CoV-2 D614G, with 50% neutralization titers ranging from <16 to 66 (Fig. 1H), and none of the donors showed detectable serum neutralization against Beta or Delta (Fig. 1I, J). Homologous ChAdOx1 booster immunization resulted in an overall 4.6-fold increase (geometric mean titer = 82) in serum neutralizing activity against SARS-CoV-2 D614G, but neutralizing titers remained at the lower limit of detection in 21% (6/28) of donors (Fig. 1H). We also detected a small but significant (1.5 to 2.4-fold) rise in serum neutralizing titers against both VOCs following homologous boost, although 40% and 50% of donors lacked detectable serum neutralization against Delta and Beta, respectively (Fig. 1I, J). In contrast to the relatively weak serum neutralizing responses observed following ChAdOx1 boost, heterologous mRNA-1273 booster vaccination significantly enhanced serum neutralizing titers to SARS-CoV-2 D614G and both VOCs (geometric mean titers = 1595, 727, 854 for D614G, Beta and Delta, respectively) and detectable serum neutralizing activity was observed in all donors against all three SARS-CoV-2 isolates (Fig. 1H-J). Stratification of nAb titers revealed that 96%, 100%, and 88% of mRNA-1273 boosted donors exhibited neutralizing titers >1:200 against SARS-CoV-2 D614G, Delta, and Beta, respectively, compared to only 21%, 10%, and 0% of donors who received a homologous ChAdOx1 boost (Fig. 1K-M). Taken together, heterologous ChAdOx1:mRNA-1273 prime-boost vaccination elicits superior binding and neutralizing antibody responses to SARS-CoV-2 and VOCs relative to homologous ChAdOx1:ChAdOx1 vaccination.

Previous studies of other viral infections have shown that booster vaccination typically induces a rapid and robust antigen-specific plasmablast response that peaks at approximately day 7 following immunization and accounts for a relatively large proportion of peripheral blood B cells (*18, 19*). We therefore assessed the frequency of plasmablasts in peripheral blood 7-10 days following homologous and heterologous booster vaccination (Fig. S2A). Although we observed modest plasmablast expansion in a subset of ChAdOx1 boosted donors, the median frequency of plasmablasts across all donors was not significantly elevated relative to pre-boost or unpaired pre-pandemic healthy donor samples (Fig. 2A). In contrast, heterologous booster immunization induced a robust plasmablast response in almost all donors. In these donors, the frequency of plasmablasts averaged 3% on day 7-10 post-boost and accounted for up to 7% of all CD19^+^ peripheral blood B cells (Fig. 2A).

**Figure 2.**
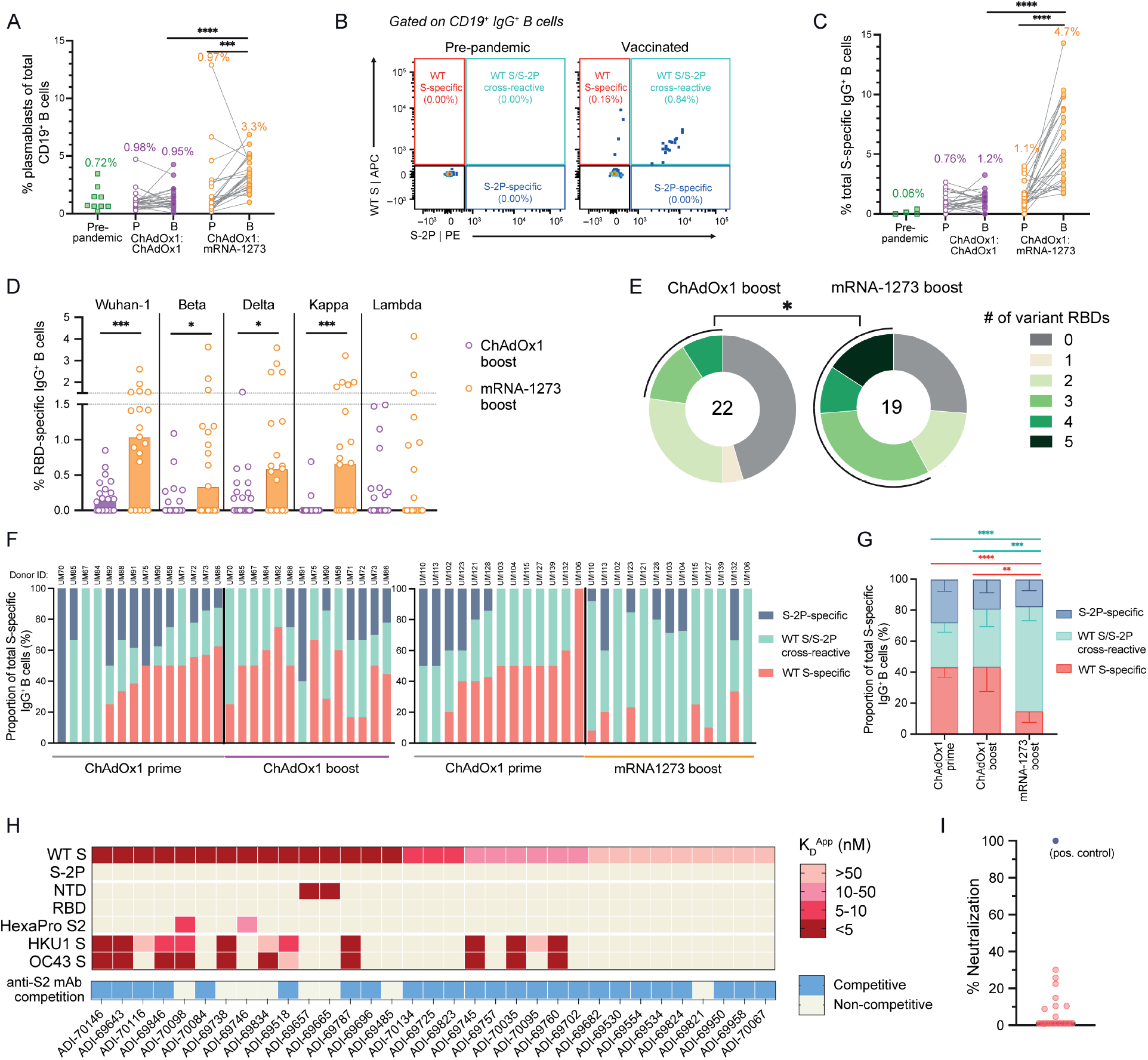
SARS-CoV-2 S-specific B cell responses induced by homologous and heterologous prime-boost vaccination. **(A)** Frequencies of plasmablasts among circulating CD19^+^ B cells following prime and boost vaccination. Plasmablasts are defined as CD19^+^CD20^—/lo^CD71^+^ cells. Pre-pandemic donor samples (N=9) were included for comparison. Median values are shown above data points. **(B)** Representative fluorescence-activated cell sorting (FACS) gating strategy used for identifying WT S-specific, S-2P-specific, and WT/S-2P cross-reactive IgG^+^ B cells. **(C)** Frequencies of total (WT + S-2P) SARS-CoV-2 S-reactive B cells among circulating IgG^+^ B cells, as determined by flow cytometry. Median frequencies are shown above data points. **(D)** Frequency of circulating IgG^+^ B cells reactive with RBDs encoding mutations present in Beta, Delta, Kappa, and Lambda variants. The height of each bar indicates median frequency. **(E)** Proportion of donors with detectable B cell reactivity with the indicated number of variant RBDs. The total number of donors analyzed is indicated in the center of the pies. Statistical significance was determined by Fisher’s exact test and calculated based on the proportion of donors with B cells displaying reactivity to ≥3 variant RBDs. **(F)** Proportions of WT S-specific, WT S/S-2P cross-reactive, and S-2P-specific B cells among total S-specific B cells following prime and homologous (left) or heterologous (right) boost immunization. Donors with S-specific B cell frequencies <1% of total IgG^+^ B cells at either time point were excluded from this analysis. Donor IDs are denoted above each bar. **(G)** Mean proportions of WT S-specific, WT S/S-2P cross-reactive, and S-2P-specific B cells across all donors within each cohort. Error bars indicate 95% confidence intervals. **(H)** Apparent binding affinities (K_D_^App^) of WT S-specific monoclonal antibodies for WT S, S-2P, prefusion S subdomains (NTD, RBD, prefusion-stabilized S2), HKU1 S, and OC43 S, as determined by biolayer interferometry (BLI). Competitive binding with an anti-S2 antibody (ADI-69962), as determined by a BLI competitive sandwich assay, is indicated below the heatmap. **(I)** Neutralizing activity of WT S-specific antibodies against MLV-SARS-CoV-2 Wuhan-1 at a concentration of 1 *µ*g/ml. A previously described anti-RBD neutralizing antibody (ADG-2) was included as a positive control and is shown in purple (*25*). Statistical comparisons between paired prime and boost samples were determined by Wilcoxon pair-matched rank sum test (A and C). Statistical comparisons between vaccination cohorts were determined by two-sided Mann-Whitney U tests (D) and two-sided Kruskal-Wallis test by ranks with subsequent Dunn’s multiple comparisons (C and G). \**P* < 0.05, \*\**P* < 0.01, \*\*\**P* < 0.001, ****P<0.0001. P, Prime; B, Boost; K_D_^App^; apparent equilibrium constant.

We next investigated the magnitude and specificities of the S-specific MBC response induced following both prime and boost immunization. Because ChAdOx1 encodes WT S, which may elicit B cell responses to both pre- and post-fusion conformations of S, we evaluated B cell responses to both prefusion-stabilized (S-2P) and WT (unstabilized) forms of the S protein by flow cytometry (Fig. 2B and Fig. S2B). Notably, SDS-PAGE analysis revealed that the recombinant WT S preparation used for B cell staining contained both uncleaved and cleaved forms of S, the latter of which likely represents a mixture of prefusion, post-fusion, and non-native or dissociated forms of S (Fig. S3). The frequency of total S (WT + S-2P)-specific IgG^+^ B cells ranged from 0-3.7% following ChAdOx1 prime immunization, and homologous ChAdOx1 booster immunization did not significantly enhance this response in most donors (Fig. 2C). In contrast, mRNA-1273 booster vaccination induced a robust expansion of S-specific IgG^+^ B cells in most donors, averaging a 4-fold increase over the corresponding ChAdOx1 prime-induced responses (Fig. 2C). Similarly, mRNA-1273 booster immunization expanded S-specific IgM^+^CD27^+^ and IgA^+^ MBC cell populations, although the frequencies were lower than those observed for IgG^+^ B cells (Fig. S4). Consistent with the responses observed for full-length S, mRNA-1273 booster vaccination elicited significantly higher magnitude B cell responses to individual subdomains within prefusion S (RBD, NTD, and prefusion-stabilized S2), including RBDs encoding mutations found in the Beta, Delta, and Kappa variants, relative to homologous ChAdOx1 boost (Fig. 2D, Fig. S5, and Fig. S6). Only 23% of ChAdOx1 boosted donors displayed detectable B cell reactivity with ≥3 variant RBDs compared to 58% of mRNA boosted donors (Fig. 2E). Overall, the results suggest that heterologous ChAdOx1:mRNA-1273 prime-boost vaccination induces a more robust B cell response to SARS-CoV-2 and VOCs relative to homologous ChAdOx1:ChAdOx1 immunization.

The use of dual-labeled SARS-CoV-2 S probes for B cell staining allowed us to assess the frequencies and proportions of WT S-specific, WT S/S-2P-reactive, and S-2P-specific B cells induced following prime and boost immunization. In the majority of donors (19/27), ≥ 30% of the total S-specific IgG^+^ MBC response induced by ChAdOx1 prime immunization was directed towards epitopes expressed only on WT S, with the remaining B cells displaying either specificity for S-2P or cross-reactivity between WT S and S-2P (Fig. 2F, G). As expected, homologous ChAdOx1 booster vaccination did not significantly modify this response in most donors (Fig. 2F, G). In contrast, we observed a massive decline in the proportion of circulating WT S-specific B cells following mRNA booster immunization in most donors (Fig. 2F, G). Correspondingly, booster immunization with mRNA-1273 preferentially expanded WT S/S-2P cross-reactive B cells; these specificities were present in peripheral blood at a median frequency of 0.3% prior to booster immunization and 3.4% following mRNA-1273 boost, while the frequency of WT S-specific cells declined from 0.2% to 0% (Fig. S7). Thus, in addition to driving a robust expansion of ChAdOx1 prime-induced B cells, heterologous mRNA-1273 booster immunization re-directs the B cell response towards epitopes expressed on the prefusion form of the S protein.

To characterize the specificities and functional properties of the WT S-specific antibodies induced by ChAdOx1 prime immunization, we cloned and expressed 33 monoclonal antibodies from single WT S-specific B cells in five donors. The antibodies utilized a diversity of variable heavy-(VH) and light-chain (VL) germline genes, and 28 out of 33 contained somatic mutations, consistent with an MBC origin (Fig. S8). The avid binding affinities of the antibodies for recombinant WT S ranged from 1 to 36 nM and none displayed detectable binding to S-2P (Fig. 2H). Over 85% (29/33) of the WT S-specific antibodies failed to bind to recombinant subdomains comprising prefusion S (RBD, NTD, and prefusion-stabilized S2) and 40% displayed reactivity with HKU1 and/or OC43 S, suggesting recognition of conserved epitopes within postfusion S2. Given the lack of availability of recombinant postfusion S antigens, we evaluated this possibility by performing competitive binding assays with an anti-S2 antibody (ADI-69962) that targets an epitope expressed on recombinant WT S, S-2P, and prefusion-stabilized S2 (Fig. S9). Seventy-five percent (25/33) of the WT S-specific antibodies showed competitive binding with ADI-69962, supporting recognition of an antigenic site within the S2 subunit that is not expressed on S-2P (Fig. 2H). Consistent with their lack of reactivity with prefusion S, none of the WT S-specific antibodies displayed >50% neutralizing activity against SARS-CoV-2 Wuhan-1 at a 1 *µ*g/ml concentration in an MLV-based pseudovirus assay (*20*) (Fig. 2I). We conclude that a relatively large proportion of the SARS-CoV-2 S-specific B cell response induced by homologous ChAdOx1 prime-boost immunization is comprised of non-neutralizing anti-S2 specificities that fail to bind prefusion S.

To determine whether homologous and heterologous booster vaccination regimens induce distinct B cell responses to prefusion S, we obtained 163 and 252 paired VH and VL sequences from single S2P-reactive B cells from 4 donors in each cohort following ChAdOx1 or mRNA-1273 booster immunization, respectively (Table S2). Both booster regimens induced highly diverse B cell responses, with 0 to 8.6% of antibodies belonging to expanded clonal lineages (Fig. S10). IGHV3-30 was significantly over-represented in the S-reactive MBC compartment in both ChAdOx1 and mRNA-1273 boosted individuals, as observed previously in antibodies isolated from naturally infected and mRNA vaccinated donors (*21, 22*) (Fig. S11A). The antibodies isolated from both donor cohorts also displayed similar levels of somatic hypermutation (SHM), which ranged from a median of 4-7 and 6-7 VH nucleotide substitutions for ChAdOx1 and mRNA-1273 boosted donors, respectively (Fig. S12A). In 7 out of 8 donors, >90% of sequences contained somatic mutations, consistent with a recall response mediated by pre-existing memory B cells induced by ChAdOx1 prime immunization (Fig. S12B).

Furthermore, the overall levels of SHM in the antibodies isolated from both donor cohorts were comparable to that observed in antibodies previously isolated from SARS-CoV-2 convalescent individuals at a similar time point following infection (approximately 3 months), suggesting that the early kinetics of affinity maturation following ChAdOx1 prime immunization may be similar to that of natural infection (Fig. S12). Overall, the results demonstrate that the genetic features of S-2P-reactive antibodies induced by homologous and heterologous booster vaccination are highly similar, with both groups rich in clonally expanded and somatically mutated sequences.

We next measured the binding affinities and neutralizing activities of the S-2P-reactive antibodies induced by homologous and heterologous prime-boost vaccination. Although the antibodies isolated from both cohorts displayed a wide range of Fab binding affinities for recombinant S-2P, those isolated from mRNA boosted donors exhibited overall higher affinities (median K_D_ = 12 nM) compared to those isolated from ChAdOx1 boosted donors (median K_D_>100 nM) (Fig 3A). This difference may be associated with expression of WT S by the ChAdOx1 vaccine, which may lead to the activation of B cells with sub-optimal affinity for S-2P. Interestingly, although a relatively large proportion of antibodies isolated from both donor cohorts bound with high affinity to S-2P, only a small subset of binding antibodies (6-13% and 8-21% for ChAdOx1 and mRNA-1273 boosted donors, respectively) displayed >50% neutralizing activity against SARS-CoV-2 Wuhan-1 at a concentration of 1 *µ*g/ml in a pseudovirus neutralization assay (Fig. 3B). Thus, the neutralizing antibody response represents only a minor fraction of the total prefusion S-reactive binding response induced by both homologous and heterologous prime-boost immunization.

**Figure 3.**
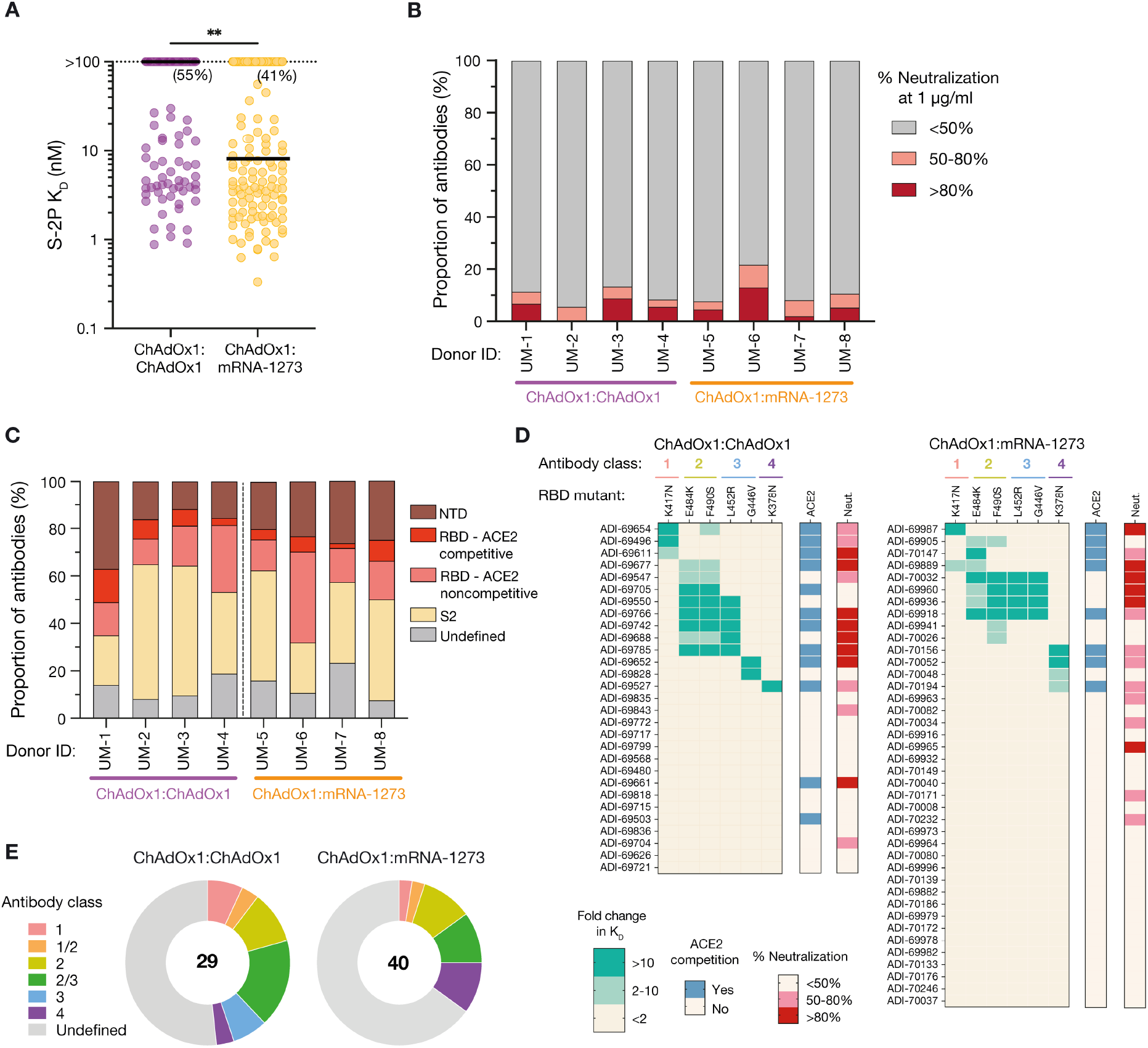
Binding and neutralization properties of monoclonal antibodies isolated from ChAdOx1 and mRNA-1273 boosted donors. **(A)** Fab binding affinities for S-2P, as determined by BLI. Antibodies with no detectable monovalent binding activity are excluded and those with weak binding affinities that could not be fit to a 1:1 binding model are plotted as K_D_ >100 nM. Black bars indicate medians. Values in parentheses indicate the percentage of antibodies with K_D_>100 nM. **(B)** Proportion of antibodies exhibiting <50%, 50-80%, and >80% neutralization activity against MLV-SARS-CoV-2 Wuhan-1 at a concentration of 1 *µ*g/ml. **(C)** Proportion of S-2P-reactive antibodies directed to each of the indicated subdomains within prefusion S. Competitive hACE2 binding was determined using a BLI-based competition assay. **(D)** Heatmaps displaying fold change in binding affinity of anti-RBD antibodies to variant RBDs containing the indicated single point mutations. Competitive hACE2 binding activity and percentage neutralization against MLV-SARS-CoV-2 Wuhan-1 at a concentration of 1 *µ*g/ml are shown in the bars on the right. **(E)** Summary of the distribution of anti-RBD antibodies belonging to each of the indicated classes. The numbers in the center of the pies indicate the total number of antibodies analyzed. Statistical comparisons were made by two-sided Mann-Whitney U tests (A). K_D,_ equilibrium dissociation constant. \*\**P* < 0.01.

To examine if and how the type of booster immunization impacts B cell immunodominance hierarchy to prefusion S, we evaluated the proportion of S-2P-reactive antibodies directed to the NTD, RBD, and prefusion-stabilized S2 subdomains in each donor from which monoclonal antibodies were isolated. Across the selected donors in both cohorts, we observed relatively similar proportions of antibodies targeting each individual subdomain within prefusion S, although S2-directed antibodies dominated the response in a subset of donors in both groups (Fig. 3C). Unlike the response to WT S, antibodies targeting conserved epitopes expressed on OC43 and HKU1 S constituted only a minor fraction of the S-2P-reactive response elicited by both ChAdOx1 and mRNA-1273 booster vaccination (5.5% and 4.8%, respectively) (Fig. S13). RBD-directed hACE2 blocking antibodies also represented a small proportion of the S-2P-reactive binding response in both donor cohorts, ranging from 3-14% and 2-9% in ChAdOx1:ChAdOx1 and ChAdOx1:mRNA-1273 immunized donors, respectively (Fig. 3C). As expected, these rare anti-RBD antibodies constituted the majority of the neutralizing response, thus explaining the limited number of neutralizing antibodies observed among total S-2P binding antibodies (Fig. S14). To further map the epitopes recognized by the RBD-directed antibodies, we evaluated their binding reactivities with recombinant RBDs containing mutations associated with escape from common classes of antibodies, including K417N (class 1), E484K and F490S (class 2), L452R and G446V (class 3), and K378N (class 4) (*23, 24*). As observed for the individual subdomains within prefusion S, both booster regimens induced comparable proportions of antibodies targeting common antigenic sites within the RBD (Fig. 3D-E). In conclusion, although homologous ChAdOx1 booster vaccination induces a higher frequency of WT S-specific antibodies compared to heterologous mRNA-1273 boost, both immunization regimens establish similar immunodominance hierarchies to prefusion-stabilized S.

Finally, we assessed the RBD- and NTD-directed antibodies isolated from both donor cohorts for reactivity with emerging SARS-CoV-2 VOCs and variants of interest (VOIs). Using a multiplexed bead-based flow cytometry assay, we determined their binding affinities for recombinant RBD and NTDs containing mutations found in the Beta, Gamma, Delta, Kappa, and Lambda VOCs/VOIs. Interestingly, 41% (12/29) of anti-RBD antibodies derived from ChAdOx1 boosted individuals displayed reduced binding activity to ≥1 VOC/VOI RBD as compared to only 20% (8/40) of antibodies isolated from mRNA-1273 boosted individuals (Fig. 4A and Fig. S15A). Correspondingly, for each VOC/VOI RBD tested, a larger proportion of antibodies (80-90%) isolated from mRNA-1273 boosted donors retained binding activity (<2-fold decrease in affinity relative to WT) compared to those derived from ChAdOx1 boosted donors (62-82%) (Fig. 4B). Furthermore, the mRNA-elicited antibodies bound to both WT and variant RBDs with significantly higher affinities (median K_D_=1.4-3.7 nM) relative to the ChAdOx1-induced antibodies (median K_D_=4.9-20.3 nM) (Fig. 4C), potentially explaining their increased breadth of binding. Consistent with these results, 75% of NTD-directed antibodies isolated from ChAdOx1 boosted donors displayed loss of binding to ≥1 VOC/VOI NTDs compared to only 53% of antibodies isolated from mRNA-1273 boosted donors (Fig. 4D-E and Fig. S15B). Similar to the RBD-directed antibodies, the NTD-specific antibodies isolated from mRNA-1273 boosted donors trended toward higher binding affinities across all three VOCs compared to antibodies derived from ChAdOx1 boosted donors (Fig. 4F). Overall, a significantly larger fraction of anti-RBD and -NTD antibodies (70%) derived from mRNA boosted donors retained reactivity with all three VOC NTDs or RBDs relative to ChAdOx1 boosted donors (51%) (Fig. 4G). Thus, heterologous mRNA-1273 immunization appears to skew the early secondary B cell response toward higher affinity clones with improved breadth of variant recognition compared to homologous ChAdOx1 immunization.

**Figure 4.**
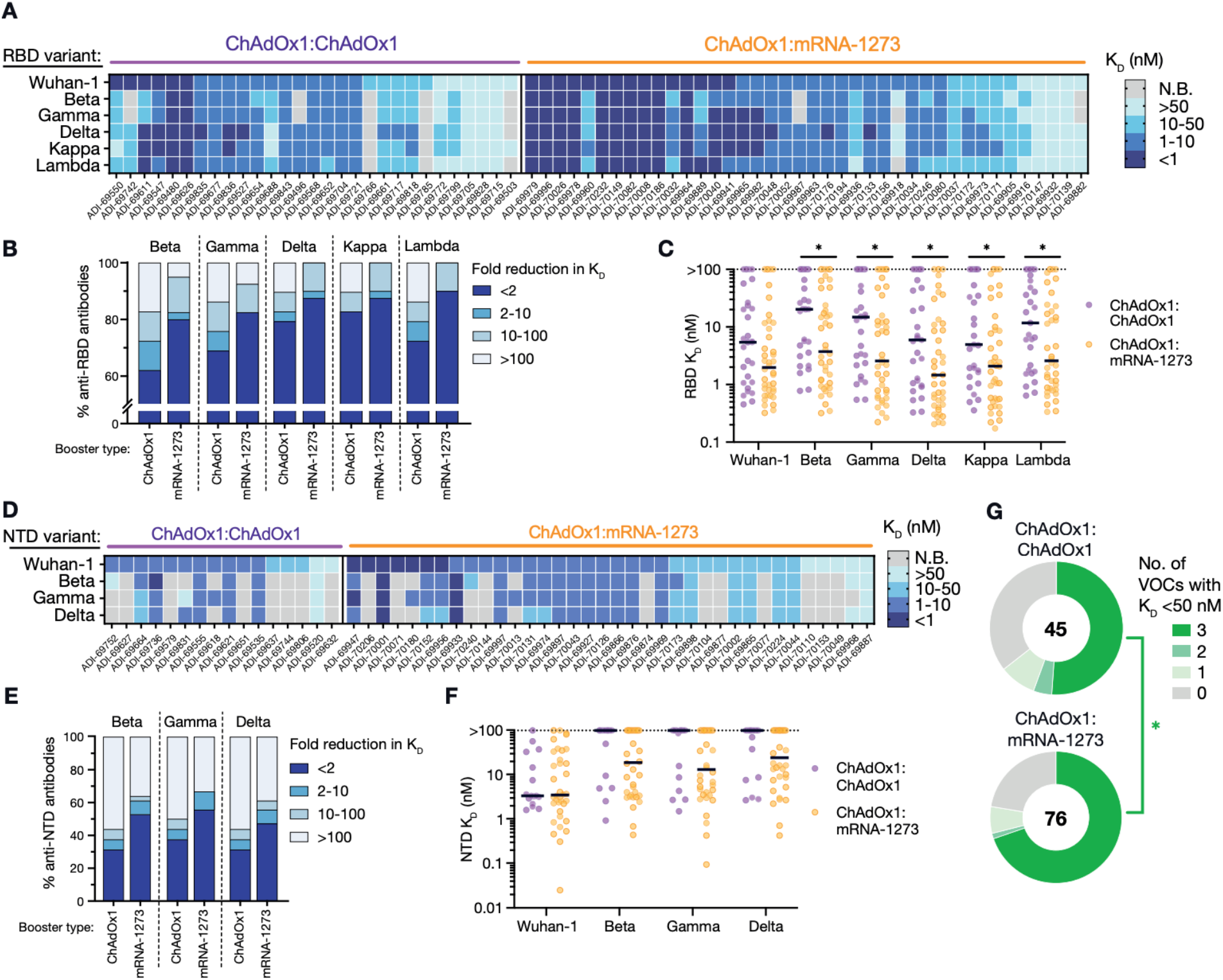
Breadth of antibody binding to circulating SARS-CoV-2 variants. **(A)** Fab binding affinities of antibodies derived from ChAdOx1 or mRNA-1273 boosted donors to the Wuhan-1 RBD and RBDs incorporating mutations present in the Beta, Gamma, Delta, Kappa, and Lambda variants, as determined using a bead-based flow cytometric assay. **(B)** Proportion of anti-RBD antibodies with the indicated fold reduction in binding affinity to variant RBDs relative to the Wuhan-1 RBD. **(C)** Fab binding affinities of anti-RBD antibodies to Wuhan-1 and variant RBDs. Antibodies that did not reach half-maximal saturation at the highest concentration tested (100 nM) are shown as K_D_ >100 nM. Black bars denote medians. **(D)** Fab binding affinities of anti-NTD antibodies to the Wuhan-1 NTD and NTDs encoding mutations present in the Beta, Gamma, and Delta variants. **(E)** Proportion of NTD-directed antibodies with the indicated fold reductions in binding activity to variant NTDs relative to the Wuhan-1 NTD. **(F)** Fab binding affinities of anti-NTD antibodies to Wuhan-1 and variant NTDs. Antibodies that did not reach half-maximal saturation at 100 nM are shown as K_D_ >100 nM. Black bars denote medians. **(G)** Proportion of antibodies with K_D_ ≤ 50 nM to the indicated number of variant RBDs or NTDs. The number in the center of the pie indicates the total number of antibodies tested. Statistical significance was determined by two-sided Mann-Whitney U tests (C and F) or Fisher’s exact test (G). N.B., non-binding; K_D,_ equilibrium dissociation constant. \**P* < 0.05.

In conclusion, heterologous ChAdOx1:mRNA-1273 prime-boost immunization induces significantly broader and more potent serum neutralizing antibody and MBC responses against WT SARS-CoV-2 and VOCs relative to homologous two-dose ChAdOx1 vaccination, and this difference appears to be driven by both the magnitude and quality of the early secondary B cell response. Expression of WT S by ChAdOx1 appears to distract the B cell response away from sites of vulnerability present on prefusion S, and homologous booster vaccination further expands these non-functional specificities. In contrast, heterologous booster immunization with mRNA-1273, which encodes S-2P, re-directs the B cell response toward epitopes expressed on prefusion-stabilized S. Furthermore, mRNA-1273 activates B cells with higher affinity for prefusion S and greater breadth of reactivity with VOCs relative to ChAdOx1. The molecular basis for this difference remains to be determined but may be associated with higher cell-surface expression of S-2P relative to WT S, differences in the presentation of S on the surface of antigen-presenting cells, and/or the distinct innate immunostimulatory properties of mRNA versus adenoviral particles. Finally, although heterologous ChAdOx1:mRNA-1273 prime-boost immunization shows superior immunogenicity relative to two-dose ChAdOx1, the B cell response induced by both vaccination regimens is dominated by non-neutralizing antibodies. Rationally designed immunogens that focus the B cell response on conserved, neutralizing epitopes may further enhance the potency, breath, and durability of protection against SARS-CoV-2, future emerging VOCs, and potentially pre-emergent SARS-like viruses.

## Data Availability

Antibody
sequences have been deposited in GenBank (#OL697893-OL698712). All other data are available in the manuscript or
supplementary materials. IgGs are available from L.M.W. under a material transfer agreement
(MTA) from Adagio Therapeutics, Inc.

## Acknowledgements

We thank study nurse Ida-Lisa Persson and personnel at the Clinical Research Center at Umeå University Hospital for support, enrollment of study subjects, and sampling. We also thank Linnea Vikström, Maj Järner and Mikaela Lagerqvist at Umeå University for processing of samples and Alicia Edin, Umeå University Hospital for the summary of clinical data. We also acknowledge Parexel for assistance with figure preparation, and E. Krauland, J. Nett, M. Vasquez, L. Connolly, and E. Hershberger for helpful comments on the manuscript. All IgGs were sequenced by Adimab’s Molecular Core and produced by the High Throughput Expression group.

## Author contributions

L.M.W. and M.N.E.F. conceived and designed the study. C.A. and J.N. are principal investigators for the clinical trial and supervised sample collection. C.I.K. performed serum and B cell analyses, single B cell sorting, and antibody characterization. E.R.C. designed and supervised developability and biolayer interferometry assays. C.I.K. and M.S. developed, designed, and performed pseudovirus neutralization assays. C.I.K. and M.E.A. developed, designed, and performed multiplexed flow cytometry assays. C.I.K., E.R.C. and L.M.W. analyzed the data. C.I.K. and L.M.W. wrote the manuscript, and all authors reviewed and edited the paper.

## Data and material availability

Antibody sequences have been deposited in GenBank. All other data are available in the manuscript or supplementary materials. IgGs are available from L.M.W. under a material transfer agreement (MTA) from Adagio Therapeutics, Inc.

## Supplementary Materials

Materials and Methods

Figures S1-S15

Table S1-S2

