## Supplementary Material for "Broad anti-SARS-CoV-2 antibody immunity induced by heterologous ChAdOx1/mRNA-1273 prime-boost vaccination"

**Materials and Methods**

**Study subjects and sample collection.**

Subjects were recruited by informed consent to participate in a 4 -year follow-up study to assess the safety and immunogenicity of COVID-19 vaccines in Sweden. Participants received one dose of ChAdOx-1 and 9 to 12 weeks later (65-87 days), were offered a booster immunization with either ChAdOx-1 nCoV-19 or mRNA-1273. Of the 55 study subjects, 28 chose to receive ChAdOx-1 whereas 27 chose mRNA-1273 as their second dose. Participants with known prior SARS-CoV-2 infection were excluded. Demographics of the two different groups are shown in Table S1. All individuals were bled for serum, plasma and PBMCs on the day of the booster injection and then again 7-10 days and 1 month after the booster injection. On each sampling occasion, venous blood was collected and PBMCs and plasma isolated in BD EDTA Vacutainer® CPT™ tubes. PBMCs were frozen in 90% fetal calf serum supplemented with 10% DMSO and stored in liquid nitrogen until use. Plasma and serum were stored at -80°C.

**Study oversight and ethical permits**

The clinical trial, CoVacc - Immune response to vaccination against Covid-19, an open multicenter phase IV study, was approved by the Swedish Ethics Review Authority (Dnr 2021-00055) and the Medical Products Agency Sweden. The study was registered at European Clinical Trials Database (EUDRACT Number 2021-000683-30) before the first patient was enrolled. Umeå University, Sweden served as trial sponsor and the Clinical Research Center, University Hospital of Northern Sweden was monitoring the study for regulatory compliance. Individuals were included after informed consent and data were stored in accordance with the EU General Data Protection Regulation.

**Recombinant antigen production.**

For production of prefusion-stabilized SARS-CoV-2 S-2P, DNA encoding residues 1-1208 of the SARS-CoV-2 spike with “PP” mutations at positions 986 and 987, “GSAS” mutations from positions 682-685 and a C-terminal T4 fibritin motif, 8X HisTag and TwinStrepTag (SARS-CoV-2 S-2P) was cloned into a pcDNA3.4 vector and transiently transfected into FreeStyle HEK 293F cells (Thermo Fisher) using polyethylenimine. Following one week of culture, the supernatants were harvested, centrifuged to remove cellular debris, and purified by Ni affinity chromatography. Recombinant protein was further purified by size exclusion chromatography using the Superose 6 column (GE Healthcare).

For production of recombinant wildtype and variant RBDs, DNA sequences of the Wuhan-1 RBD (spike residues 319-537); RBDs encoding single mutations K417N, E484K, F490S, L452R, G446V, K378N; and RBDs encoding mutations present in Beta (K417N, E484K, N501Y), Gamma (K417T, E484K, N501Y), Delta (L452R, T478K), Kappa (L452R, E484Q), and Lambda (L452Q, F490S) variants were ordered as gBlocks (IDT). RBD DNA fragments were cloned into a yeast surface display expression vector encoding an N-terminal hemagglutinin epitope tag and a C-terminal Gly4Ser linker, Avitag sequence, HRV3C protease recognition sequence, and Aga-2p. Plasmids were transformed into *S. cerevisiae* (EBY100) using the Frozen-EZ Yeast Transformation II Kit (Zymo Research) per manufacturer's directions. Transformed yeast cells were grown in SDCAA media at 30 ˚C until cell densities reached 1.0 OD_600_/ml. Cells were next transferred to SGCAA media and incubated for 16-20 hrs at 20 ˚C to induce RBD expression. Yeast surface-expressed RBDs were biotinylated using the BirA protein ligase reaction kit (Avidity) following manufacturer's recommendations and then washed three times to remove excess biotin. To cleave recombinant biotinylated RBDs from the cell surface, 20 µl (20 U) of HRV 3C protease was diluted in 1X HRV 3C cleavage buffer (Takara, Cat #7360) and incubated with 20 OD_600_ of yeast cells in a 500 µl volume for 16 hrs at 4 ˚C with end-over-end rotation. Cells were subsequently pelleted by centrifugation, and supernatant containing biotinylated RBD was aspirated and stored at -80 ˚C.

**Serum ELISAs.**

96-well half-area plates (Corning) were coated with 25 µl of recombinant antigen diluted in PBS and incubated overnight at 4 °C. Recombinant SARS-CoV-2 RBD (Sino Biological, Cat #40592-V08B), NTD (Acro Biosystems, Cat #S1D-52H6), Hexapro-stabilized S2 (Acro Biosystems, Cat #S2N-C52H5), HKU1 S (Sino Biological, Cat #40606-V08B), and OC43 S (Sino Biological, Cat #40607-V08B) antigens were coated at a concentration of 5 µg/ml. To assess the SARS-CoV-2 S-reactive response, plates were coated with a mixture of recombinant S-2P and WT S (Sino Biological, Cat #40589-V08B1) at 2.5 µg/ml concentrations of each antigen. Following antigen adsorption to plate surfaces, wells were washed with wash buffer (1X PBS with 0.05% Tween-20) and blocked with 50 µl 3% bovine serum albumin (BSA) in 1X PBS for 1 h at 37 °C. Wells were incubated with serial dilutions of human serum ranging from 1:20 to 1:655,000 in a solution of 0.1% BSA, 0.01% Tween-20 in 1X PBS for 1 h at 37 °C. Plates were then washed three times before incubating with a 1:4000 dilution of anti-human IgG horseradish peroxidase (Jackson Immunoresearch Laboratories) in 0.1% BSA and 0.01% Tween-20 in 1X PBS for 30 min at 37 °C. Plates were then washed three times to remove excess secondary reagent and developed with 25 µl of room temperature-equilibrated 1-Step™ Ultra TMB Substrate Solution (Thermo Fisher Scientific) for 5 min. The developing reaction was terminated by addition of 25 µl 2 M sulfuric acid. Absorbance was measured at 450 nm using a Spectramax microplate reader (Molecular Devices). Absorbance values were normalized by subtracting background signals from a no-serum control on each plate prior to calculating area under the curve in Graphpad Prism (version 9).

**Authentic SARS-CoV-2 plaque reduction neutralization assay.**

Serum neutralizing titers were determined for authentic SARS-CoV-2 D614G (SARS-CoV-2 strain hCoV-19/Germany/BavPat1/2020) and Beta/B.1.351 (SARS-CoV-2 strain hCoV-19/South Africa/KRISP-K005325/2020) in a plaque reduction neutralization test (PRNT) at Viroclinics Biosciences (Rotterdam, NLD). The Beta isolate contained the following mutations in the spike protein: L18F, D80A, D215G, K417N, E484K, N501Y, D614G, Q677H, R682W, A701V. Vero-E6 cells were seeded in 96-well plates overnight. SARS-CoV-2 viral particles were pre-incubated with serial dilutions of heat-inactivated sera for one h at 37 ˚C and then added to a confluent monolayer of Vero-E6 cells. After one h of incubation, cell culture supernatants were aspirated and exchanged with fresh infection medium. Cells were subsequently incubated for 16-24 hrs at 37 ˚C with 5% carbon dioxide. Next, cells were washed with DPBS, fixed with 10% formalin for 15 min at room temperature, and permeabilized with 70% ethanol. Viral infection was detected by VirSpot immunostaining, with primary detection using an anti-nucleocapsid protein antibody followed by secondary detection with peroxidase-conjugated anti-human IgG and KPL TrueBlue substrate. Blue precipitates, which indicate the presence of viral N protein, were quantified by a CTL ImmunoSpot analyzer to determine the 50% serum neutralizing titers, as described previously (*26*).

**Authentic SARS-CoV-2 CPE microneutralization assay.**

Serum neutralizing titers against a clinical isolate of the SARS-CoV-2 Delta/B.1.617.2 virus (isolate MEX-BC15/2021), containing spike mutations T19R, G142D, E156G, F157del, R158del, A222V, L452R, T478K, D614G, P681R, and D950N, were determined in a cytopathic effect (CPE)-based microneutralization assay at Retrovirox (San Diego, CA). Vero-E6 cells were seeded into 96-well tissue culture plates 24 hrs prior to the assay. Virus was incubated with 3-fold serial dilutions of heat-inactivated serum samples ranging from 1:20 to 1:43,740 in DMEM supplemented with 2% FBS for 1 h at 37 ˚C. The antibody-virus mixture was added to Vero-E6 cells and incubated for 96 h. Cell viability was measured by staining cells with 0.017% neutral red for 3 hrs. Inhibition of CPE-induced cell death was used as an indicator of neutralization activity. Following incubation, extra neutral red stain was washed away, and remaining neutral red that was retained by cellular lysosomes was extracted using a solution of 50% ethanol and 1% acetic acid. The amount of neutral red was quantified by absorbance at 450 nm using a spectrophotometer. To calculate percentage neutralization, the average absorbance of cells infected in the presence of serum dilutions was subtracted by the absorbance of cells infected in the absence of serum and then normalized to cells that were not infected with virus. To calculate the 50% neutralization titer, neutralization curves were fit by non-linear regression using a four-parameter sigmoidal dose-response model in GraphPad Prism (version 9).

**Generation of SARS-CoV-2 S-pseudotyped MLV.**

Single-cycle infection pseudoviruses were generated as previously described (*20*). Briefly, HEK293T cells were co-transfected with the following plasmids: 1) pCDNA3.3 encoding SARS-CoV-2 (NC_045512) spike genes with 19-residue C-terminal truncations, 2) MLV-based luciferase reporter gene plasmid (Vector Builder), and 3) MLV gag/pol (Vector Builder). 0.5 µg of the SARS-CoV-2 plasmid and 2 µg each of MLV luciferase and gag/pol plasmids were mixed per well and transfected using Lipofectamine 2000 (ThermoFisher Scientific) in a 6-well plate following manufacturer's protocol. Cell culture media was exchanged at 16 hrs post-transfection with fresh DMEM containing 10% FBS. Supernatants containing SARS-CoV-2 S-pseudotyped MLV particles were collected 48 hrs post-transfection, aliquoted, and frozen at -80 °C for neutralization assays.

**SARS-CoV-2 MLV pseudovirus neutralization assay.**

Neutralizing activity of monoclonal antibodies was evaluated in an MLV-based pseudovirus assay. HeLa-hACE2 reporter cells (BPS Bioscience Cat #79958) were seeded overnight at 10,000 to 15,000 cells per well in 96-well tissue culture plates (Corning). For screening of antibody neutralization activity at a single concentration, 25 µl of SARS-CoV-2 S-pseudotyped MLV particles were added to IgG in MEM/EBSS supplemented with 10% FBS to achieve a final antibody concentration of 1 µg/ml in a 100 µl volume. For determination of 50% inhibitory concentrations, MLV particles were mixed with serial dilutions of antibodies ranging from 10 µg/ml to 0.02 ng/ml. The mixture of viral particles and antibodies was incubated for 1 h at 37 ˚C with 5% carbon dioxide. Separately, HeLa-hACE2 cells were washed three times with DPBS. The virus-antibody solution was subsequently incubated with HeLa-hACE2 cells for 72 hours, after which cells were lysed with Luciferase Cell Culture Lysis 5× reagent (Promega). Luciferase activity was measured using the Luciferase Assay System (Promega) following manufacturer's directions, and relative luminescence units (RLUs) were quantified on a luminometer (Perkin Elmer). Percentage neutralization was calculated as 100*(1–[RLU_sample_– RLU_background_]/[ RLU_isotype control mAb_–RLU_background_]), and the 50% neutralization concentration was interpolated from four-parameter non-linear regression fitted curves in GraphPad Prism.

**Detection and single-cell sorting of antigen-specific B cells.**

Antigen-specific B cells were detected using mixtures of recombinant protein probes tetramerized by streptavidin (SA) conjugated to different fluorophores. All recombinant proteins were biotinylated with EZ-Link Sulfo-NHS-LC-Biotin (Thermo Fisher) at a molar ratio of seven biotin to 1 protein, and excess biotin was removed by Zebra Spin Desalting Columns. Biotinylated antigens were mixed with AlexaFluor 633-conjugated streptavidin (SA) (SA-633; Invitrogen) or PE-conjugated SA (SA-PE; Invitrogen) at a 4:1 molar ratio of antigen to SA for 30 min at 4 ˚C. For B cell specificity mapping, tetramers of WT S (Sino Biological, Cat #40589-V08B1) with SA-633 and S-2P with SA-PE were prepared individually and mixed after multimerization. For analysis of B cell reactivity with spike subdomains, recombinant SARS-CoV-2 Wuhan-1 RBD (Acro Biosystems, Cat #SPD-C52H3), NTD (Acro Biosystems, Cat #S1D-52H6), and HexaPro-stabilized S2 (Acro Biosystems, Cat #S2N-C52H5) were each tetramerized with both SA-633 and SA-PE. To determine the frequencies of B cells reactive to RBDs of circulating variants, B cells were co-stained with SA-633 tetramers of Wuhan-1 RBD and PE-conjugated tetramers of recombinant RBDs encoding mutations found in the Beta (Acro Biosystems, Cat #SPD-C52Hp), Gamma (Acro Biosystems, Cat #SPD-C52Hr), Delta (Acro Biosystems, Cat #SPD-C52Hh), Kappa (Acro Biosystems, Cat #SPD-C52Hv), or Lambda (Sino Biological, Cat #40592-V08H113) variants.

PBMCs were stained for 15 min on ice with a mixture of the following anti-human antibodies: anti-CD19 (PE-Cy7; Biolegend), anti-CD20 (BV711; Biolegend) anti-CD3 (PerCP-Cy5.5; Biolegend), anti-CD8 (PerCP-Cy5.5; Biolegend), anti-CD14 (PerCP-Cy5.5; Invitrogen), anti-CD16 (PerCP-Cy5.5; Biolegend), and a freshly prepared mixture of PE- and APC-labelled SARS-CoV-2 S protein tetramers (25 nM each). Cells were washed once with 2% BSA/1 mM EDTA in 1X PBS. Next, cells were resuspended in a mixture of propidium iodide and anti-IgM (BV421; BD Biosciences), anti-IgG (BV605; BD Biosciences), anti-IgA (FITC; Abcam), anti-CD27 (BV510; BD Biosciences), and anti-CD71 (APC-Cy7; Biolegend) and incubated for 15 min on ice. Following washing, samples were analyzed using a BD FACS Aria II (BD Biosciences). For analysis of B cell reactivity with variant RBDs, PBMCs were stained using anti-CD19 (APC-Cy7; Biolegend), anti-CD20 (APC-Cy7; Biolegend), anti-CD3 (PerCP-Cy5.5; Biolegend), anti-CD8 (PerCP-Cy5.5; Biolegend), anti-CD14 (PerCP-Cy5.5; Invitrogen), anti-CD16 (PerCP-Cy5.5; Biolegend), propidium iodide, anti-IgM (BV421; BD Biosciences), anti-IgG (BV605; BD Biosciences), and anti-IgA (FITC; Abcam). Samples were analyzed using a FACS Celesta (BD Biosciences). Class-switched B cells, defined as CD19^+^CD3^−^CD8^−^CD14^−^CD16^−^PI^−^IgM^−^ and IgG^+^ or IgA^+^, that specifically bind to WT S or cross-react with S-2P were single-cell index sorted using a BD FACS Aria II (BD Biosciences) into 96-well polystyrene microplates (Corning) containing 20 µl per well of lysis buffer [5 µl of 5X first strand SSIV cDNA buffer (Invitrogen), 1.25 µl dithiothreitol (Invitrogen), 0.625 µl of NP-40 (Thermo Scientific), 0.25 µl RNaseOUT (Invitrogen), and 12.8 µl dH2O]. Plates were centrifuged at 1000 x *g* for 20 seconds and frozen at -80 ˚C.

**Amplification and cloning of antibody variable genes.**

Antibody variable gene transcripts (VH, Vκ, Vλ) were amplified by RT-PCR using SuperScript IV enzyme (ThermoFisher Scientific), followed by two cycles of nested PCRs, as described previously (*27*). The second round of nested PCR contained 40 base pairs of 5’ and 3’ homology to restriction enzyme-digested *S. cerevisiae* expression vectors to enable homologous recombination during transformation. Briefly, amplified variable gene transcripts were chemically transformed into competent yeast cells using the lithium acetate method and plated on selective amino acid drop-out media (*28*). Individual yeast colonies were subsequently picked for sequencing and characterization.

**Expression and purification of IgGs and Fabs.**

IgGs were expressed in *S. cerevisiae* cultures grown in 24-well plates, as described previously (*27*). Briefly, after 6 days of cell culture growth, the IgG-containing supernatant was harvested by centrifugation. IgGs were purified by protein A-affinity chromatography, eluted with 200 mM acetic acid/50 mM NaCl (pH 3.5), and pH-neutralized with 1/8^th^ volume of 2 M Hepes (pH 8.0)

Fab fragments were generated by digestion of IgG with papain for 2 hrs at 30 °C before terminating the reaction with iodoacetamide. The digested mixture of Fab and Fc was subsequently purified first by Protein A agarose to remove Fc fragments and undigested IgG. The column flow-through was subsequently purified using CaptureSelect™ IgG-CH1 affinity resin (ThermoFisher Scientific). Fabs bound to the resin surface were eluted with 200 mM acetic acid/50 mM NaCl (pH 3.5) and neutralized by 1/8th volume 2 M Hepes (pH 8.0).

**Multiplexed Fab binding to SARS-CoV-2 variant RBD and NTD.**

Sphero^TM^ streptavidin-coated yellow and pink beads (Spherotech, Cat #SVFA-2552-6K, #SVFA-2558-6K) were washed out of storage solution by centrifuging for 3 min at 1000 x *g* and resuspending in a solution of 0.1% BSA, 0.05% Tween-20 in 1X PBS. For RBD antigens, 100 µl of beads were added to 500 µl of supernatant containing yeast-produced biotinylated RBD protein and incubated for one hour at room temperature. For NTD antigens, recombinant Wuhan-1 and VOC NTD antigens were obtained commercially from Acro Biosystems (Wuhan-1, Cat #S1D-C52H6; Beta, Cat #S1D-C52Hc; Gamma, Cat #S1D-C52He; Delta, Cat #S1D-C52Hh) and biotinylated using the EZ-Link Sulfo-LC-NHS-Biotinylation kit (Thermo Fisher). 100 µg of beads were mixed with 2 µg of biotinylated NTDs for one hour at room temperature. Different RBD and NTD antigens were conjugated to uniquely colored beads to enable multiplexing of beads. Following incubation, bead solutions were washed two times and resuspended in 100 µl of 0.1% BSA, 0.05% Tween-20 in 1X PBS.

In 96-well V-bottom polypropylene plates (Greiner), 0.2 µg of each colored bead coated in variant RBD or NTD were mixed together and added to each well. Beads were pelleted by centrifugation at 1000 x *g* for 3 min, and the supernatant was removed. Antibody Fab fragments were serially titrated from concentrations of 100 nM to 0.12 nM across 9 dilution steps in 1X PBS with 0.1% BSA. Beads were resuspended in 100 µl of diluted Fab samples and incubated for one hour at room temperature with orbital shaking. Next, samples were centrifuged at 1000 x *g* for 3 min and washed with 1X PBS with 0.1% BSA. Beads were subsequently incubated with Alexa Fluor 647-conjugated anti-human F(ab)'2 (Jackson Immunoresearch Laboratories, Cat # 109-606-006) at a 1:500 dilution for 20 min on ice and washed twice before analyzing on the flow cytometer (BD Biosciences FACS Canto).

Binding signals to different recombinant RBD and NTD antigens were deconvoluted by gating on each distinct bead population, as distinguished by PE and FITC signals, in FlowJo software. The median APC fluorescence intensity was calculated for each bead population and Fab concentrations tested. Fab K_D_ was derived from nonlinear regression fitting of median intensities using a four-parameter variable slope model in GraphPad Prism (version 9). Low affinity Fabs with incomplete binding curves were denoted as K_D_ of >100 nM, and a numerical value of 100 was assigned for statistical analyses. The fold reduction in K_D_ was calculated by dividing the K_D_ of variant RBD by the K_D_ of Wuhan-1 RBD.

**Bio-Layer Interferometry Kinetic Measurements (BLI).**

Apparent binding affinities were measured by biolayer interferometry (BLI) using a FortéBio Octet HTX instrument (Sartorius). All steps were performed at 25 ^o^C and at an orbital shaking speed of 1000 rpm. All reagents were formulated in PBSF buffer (PBS with 0.1% w/v BSA). For experiments with IgGs immobilized on biosensors, the IgGs (100 nM) were captured (0.6 – 1.2 nm) onto anti-human IgG (AHC) biosensors (Sartorius) and then allowed to incubate in PBSF for a minimum of 30 min. For experiments using Fc-fusion antigens (i.e. RBD-Fc or NTD-Fc), the IgG-loaded sensors were soaked (20 min) in an irrelevant IgG1 solution (0.5 mg/ml) to block remaining Fc binding sites. For experiments using strep-tagged antigens, the IgG-loaded sensors were soaked (20 min) in a biocytin solution (100 µM) to saturate remaining streptavidin binding sites. After a short (60 s) baseline step in PBSF, the IgG-loaded biosensors were exposed (180 s) to the antigen (100 nM) and then dipped (180 s) into PBSF to measure any dissociation of the antigen from the biosensor surface. Data for which binding responses were >0.1 nm were aligned, inter-step corrected (to the association step) and fit to a 1:1 binding model using the FortéBio Data Analysis Software, version 11.1.

Experiments with IgGs or Fabs in solution were performed using biotinylated or Fc-fusion proteins. Recombinant biotinylated antigens (100 nM) were immobilized (1.0 – 2.0 nm) on streptavidin biosensors (Sartorius). Fc-fusion antigens (100 nM) were captured (1.0 – 2.0 nm) to AHC biosensors (Sartorius), and then soaked (20 min) in an irrelevant IgG1 solution (0.5 mg/ml) to block remaining Fc binding sites (only for IgG in solution experiments). All sensors were then allowed to incubate in PBSF for a minimum of 30 min. After a short (60 s) baseline step in PBSF, the antigen-loaded biosensors were exposed (180 s) to the IgG or Fab (100 nM) and then dipped (180 s) into PBSF to measure any dissociation of the IgGs from the biosensor surface. Data for which binding responses were >0.1 nm were aligned and fitted as described above.

**Competitive binding analysis using BLI.**

Antibody competition with hACE2 for binding to SARS-CoV-2 RBD and with the anti-S2 antibody ADI-69962 for binding to recombinant SARS-CoV-2 WT S was evaluated by BLI using a ForteBio Octet HTX (Sartorius). All binding steps were performed at 25 ^o^C and at an orbital shaking speed of 1000 rpm. All reagents were formulated in 1X PBS with 0.1% w/v BSA.

For ACE2 competition, IgGs (100 nM) were captured to anti-human IgG capture (AHC) biosensors (Molecular Devices) to a sensor response of 1.0 nm-1.4 nm. IgG-loaded sensors were soaked (20 min) in an irrelevant IgG1 solution (0.5 mg/ml) to block remaining Fc binding sites and were allowed to equilibrate in PBSF for a minimum of 30 min. To assess any cross interactions between proteins on the sensor surface and the secondary molecules, the loaded and blocked sensors were exposed (90 s) to hACE2 receptor (300 nM) prior to the binning analysis. After a short baseline-step (60 s), IgG-loaded biosensor tips were exposed (180 s) to the recombinant SARS-CoV-2 RBD (100 nM) and then exposed (180 s) to recombinant hACE2 (300 nM).

For ADI-69962 (anti-S2 antibody) competition, ADI-69962 IgG (100 nM) was captured to anti-human IgG biosensors to a sensor response of 1.0 nm-1.4 nm. IgG-loaded sensors were soaked (20 min) in an irrelevant IgG1 solution (0.5 mg/ml) to block remaining Fc binding sites and were allowed to equilibrate in PBSF for a minimum of 30 min. Loaded and blocked sensors were exposed (90 s) to each sample antibody (300 nM) to assess any non-specific cross interactions between the sample IgG and ADI-69962 before exposure (180 s) to SARS-CoV-2 WT S (100 nM). Since WT S exists in a trimeric form, tips were exposed (180 s) to ADI-69962 (100 nM) in solution to block any remaining accessible epitopes present on protomers not directly bound to ADI-69962 loaded on the biosensor. Lastly, ADI-69962 and WT S-loaded tips were exposed (180 s) to sample IgG (100 nM). The data was y-axis normalized, and interstep corrected using the FortéBio Data Analysis Software version 11.0. Additional binding by the secondary molecule indicates an unoccupied epitope (non-competitor), while no binding indicates epitope blocking (competitor).

**Supplementary Figures**
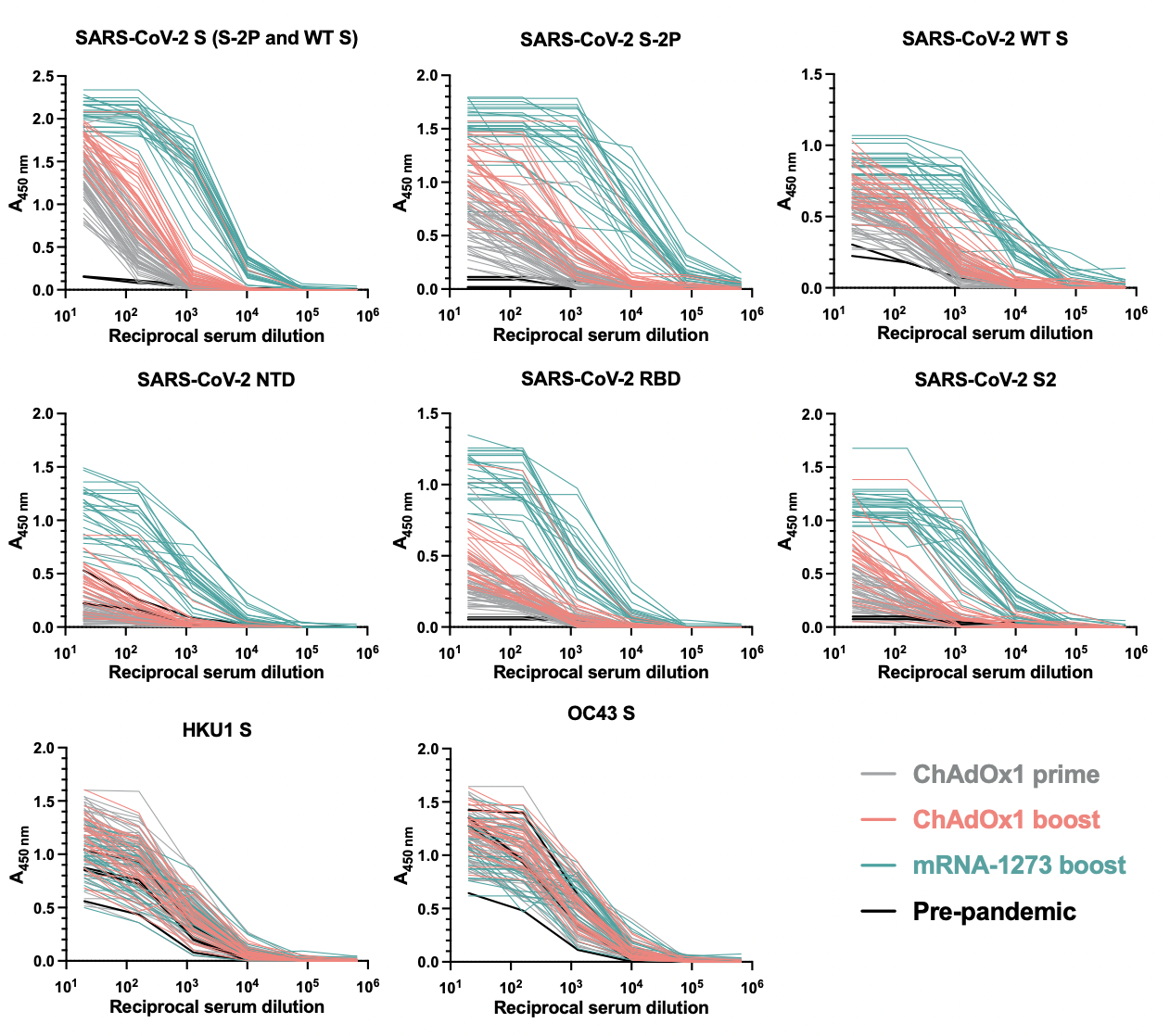

**Fig. S1.** Serum IgG binding to SARS-CoV-2, HKU1, and OC43 S antigens, as assessed by ELISA. Pre-pandemic samples were included as negative controls to assess baseline reactivities.

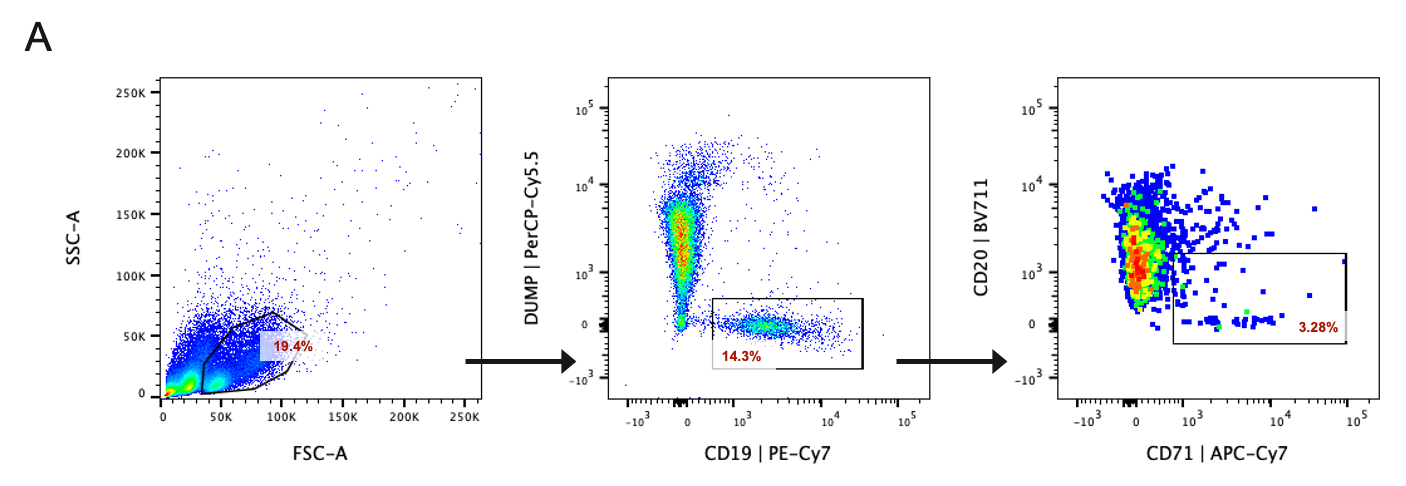

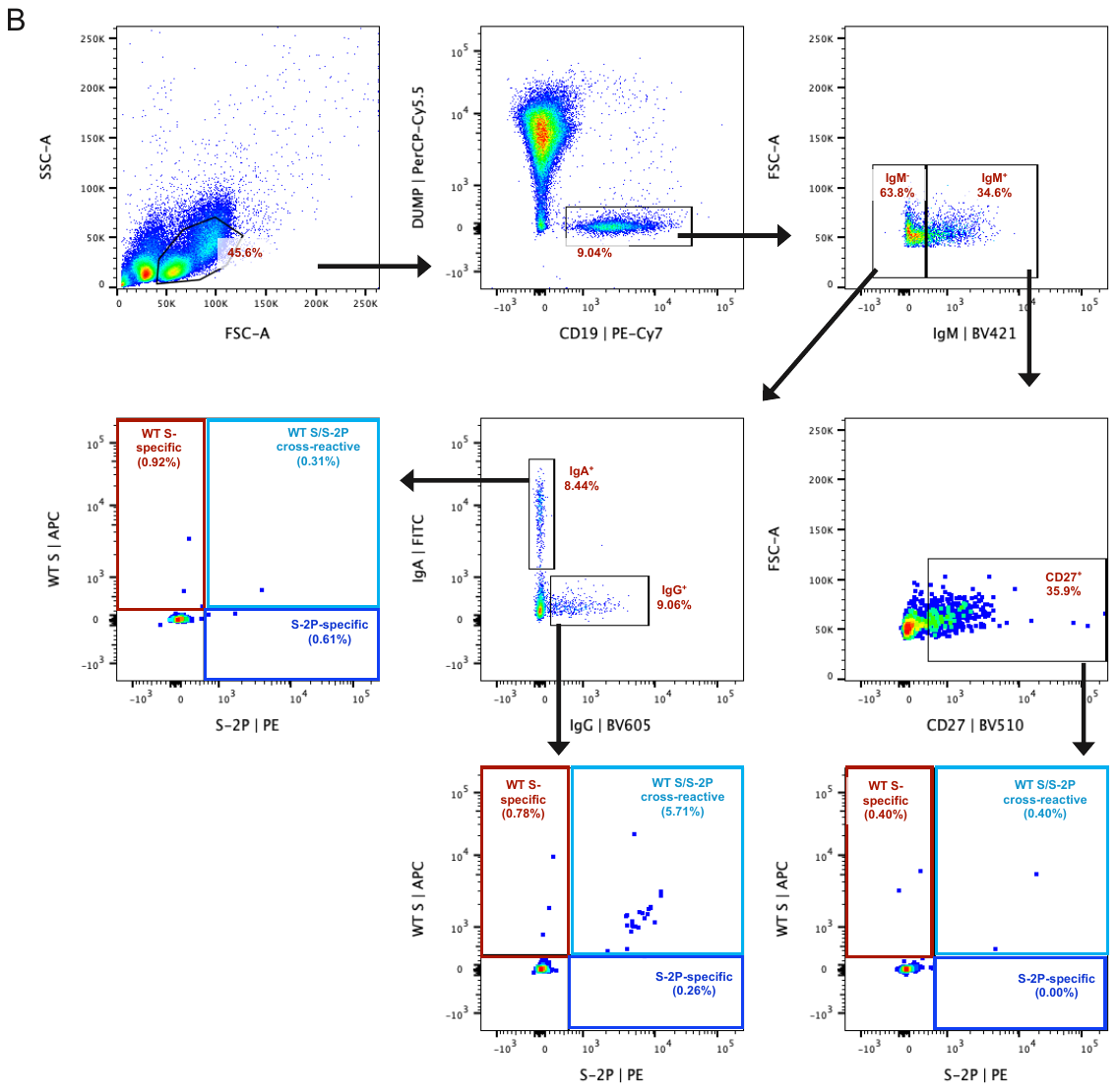

**Fig. S2.** Plasmablast and antigen-specific memory B cell staining. **(A)** Representative FACS gating strategy to determine plasmablast frequencies. Plasmablasts were identified as CD20^-/lo^CD71^+^ cells among PI^–^CD3^–^CD8^–^CD14^–^CD16^–^CD19^+^ B cells. **(B)** Representative FACS gating strategy to determine frequencies of SARS-CoV-2 S-reactive B cells among IgG^+^, IgA^+^, and CD27^+^IgM^+^ CD19^+^ B cells. SSC-A, side scatter area; FSC-A, forward scatter area; FSC-H, forward scatter height.

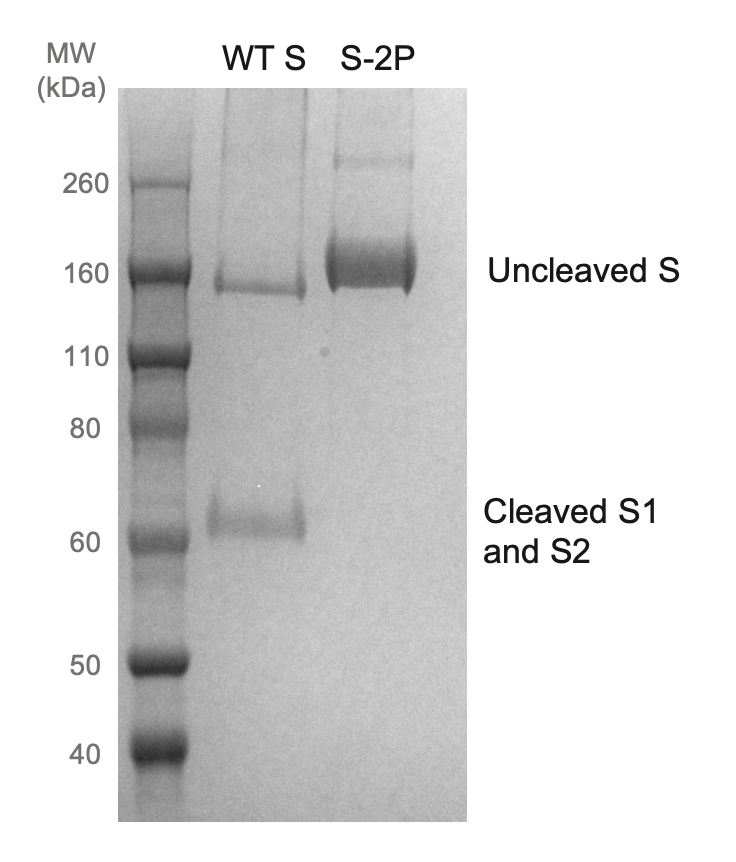

**Fig. S3.** Characterization of recombinant SARS-CoV-2 S-2P and WT S antigens via non-reducing SDS-PAGE.

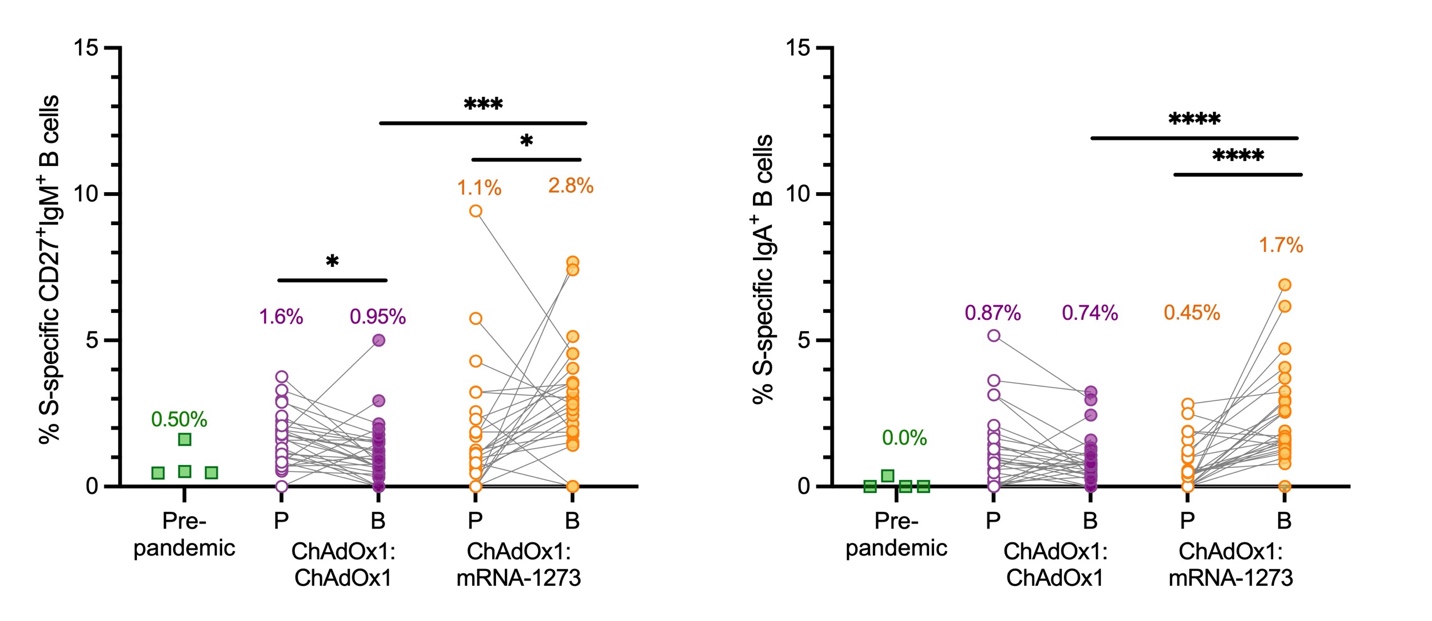

**Fig. S4.** Frequencies of total SARS-CoV-2 S-specific B cells among circulating CD27^+^IgM^+^ (left) and IgA^+^ (right) B cells, as assessed by flow cytometry. Four pre-pandemic donor samples were included for comparison. Median frequencies are shown above data points. Statistical comparisons between paired prime and boost samples were determined by Wilcoxon pair-matched rank sum test. Statistical comparisons across groups were determined by two-sided Kruskal-Wallis test by ranks with subsequent Dunn's multiple comparisons. **P* < 0.05, ****P* < 0.001, ****P<0.0001. P, Prime; B, Boost.

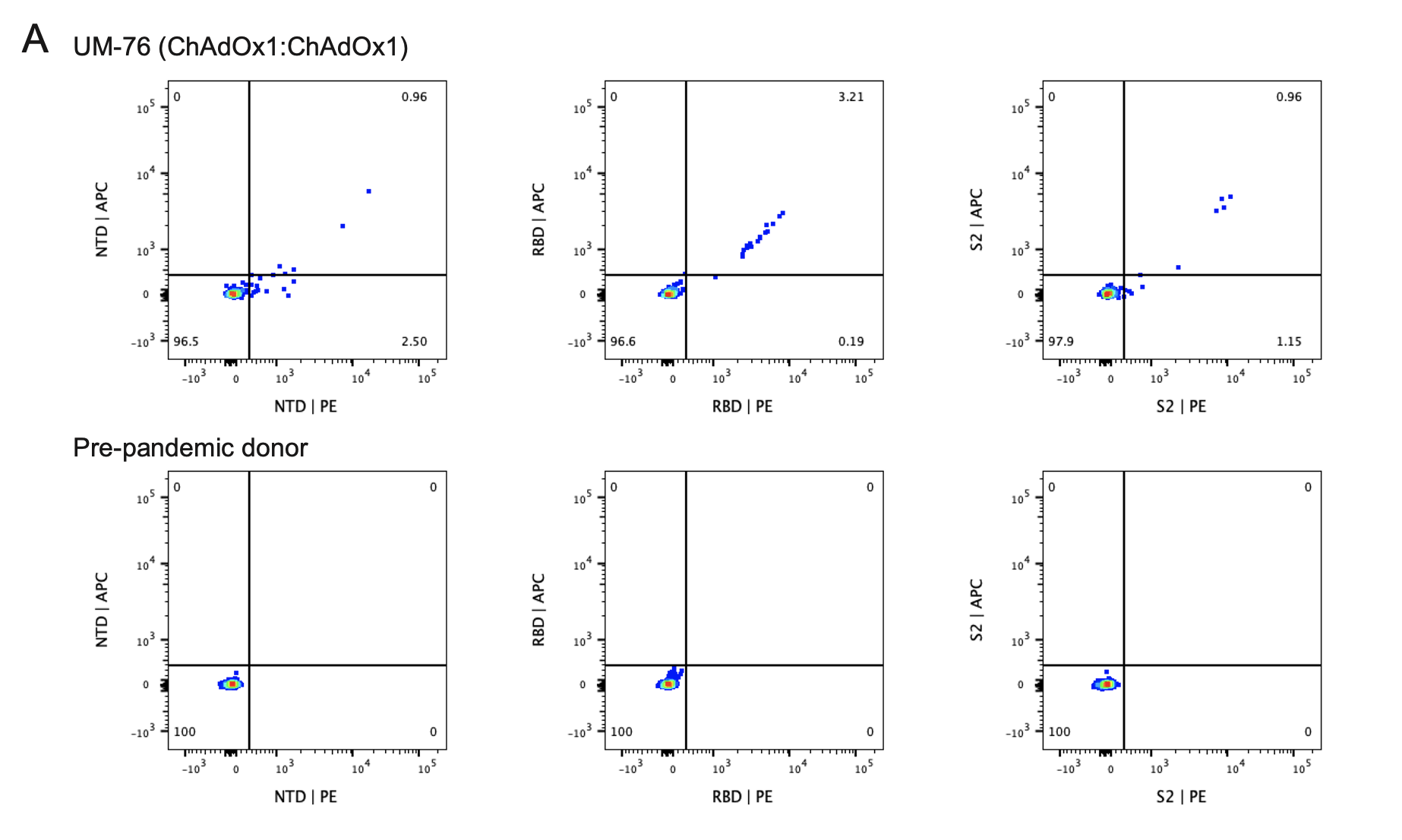

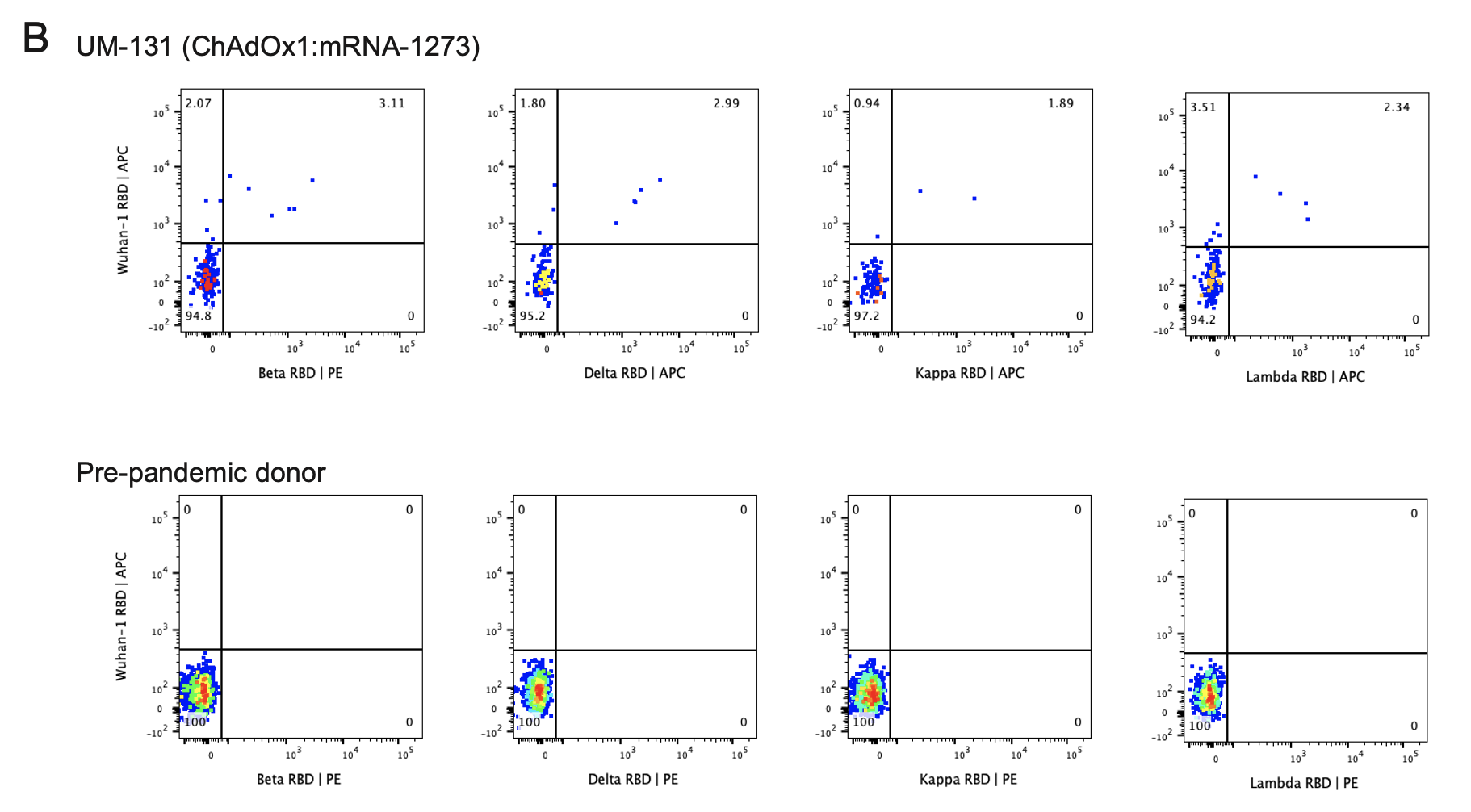

**Fig. S5.** B cell reactivity with SARS-CoV-2 S subdomains and variant RBDs. **(A)** Representative FACS plots showing B cell reactivity with recombinant NTD, RBD, and S2 subdomains among PI^–^CD3^–^CD8^–^CD14^–^CD16^–^CD19^+^IgM^–^IgG^+^ B cells. **(B)** Representative FACS plots showing B cell reactivity with recombinant Beta, Delta, Kappa, and Lambda RBDs among PI^–^CD3^–^CD8^–^CD14^–^CD16^–^CD19^+^IgM^–^IgG^+^ B cells. Pre-pandemic samples are shown for comparison.

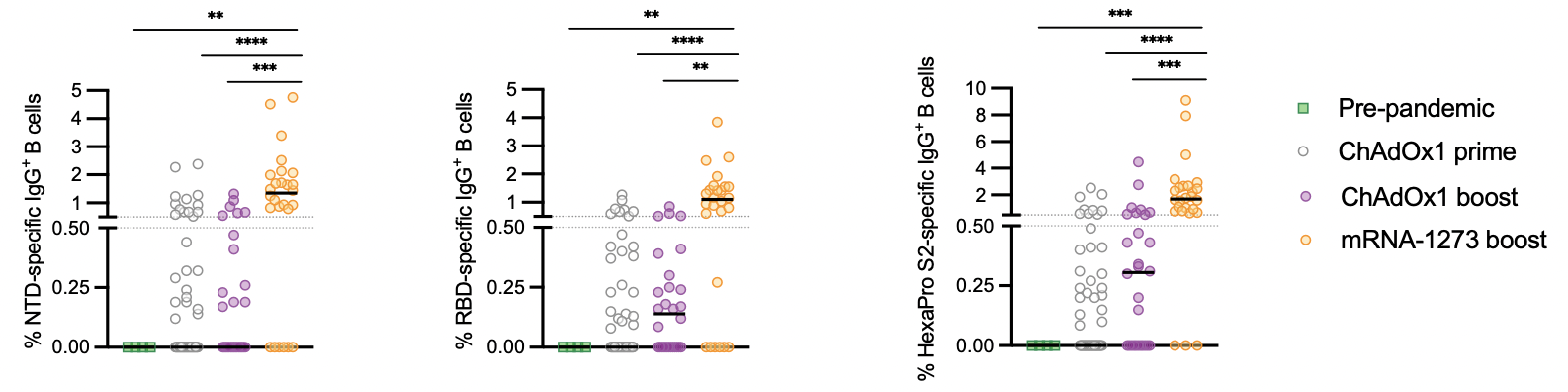

**Fig. S6.** Percentage of circulating IgG^+^ B cells reactive with recombinant NTD (left), RBD (middle), or prefusion-stabilized S2 (right) subdomains. The ChAdOx1 prime immunization group includes all donors from both cohorts. Black bars indicate medians. Statistical comparisons were determined by two-sided Kruskal-Wallis test by ranks with subsequent Dunn's multiple comparisons. ***P* < 0.01, ****P* < 0.001, ****P<0.0001.

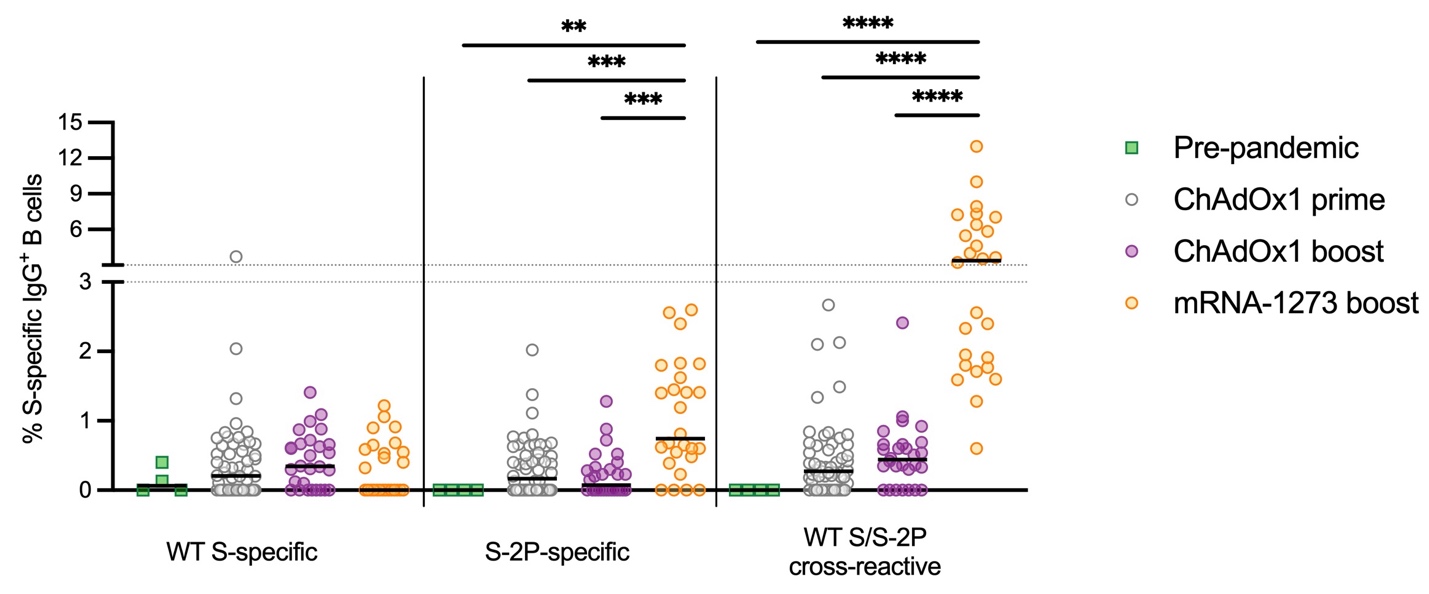

**Fig. S7.** Frequencies of WT S-specific, WT S/S-2P cross-reactive, and S-2P-specific B cells among circulating IgG^+^ B cells, as determined by flow cytometry. Black bars indicate medians. Statistical comparisons were determined by two-sided Kruskal-Wallis test by ranks with subsequent Dunn's multiple comparisons. ***P* < 0.01, ****P* < 0.001, ****P<0.0001.

.

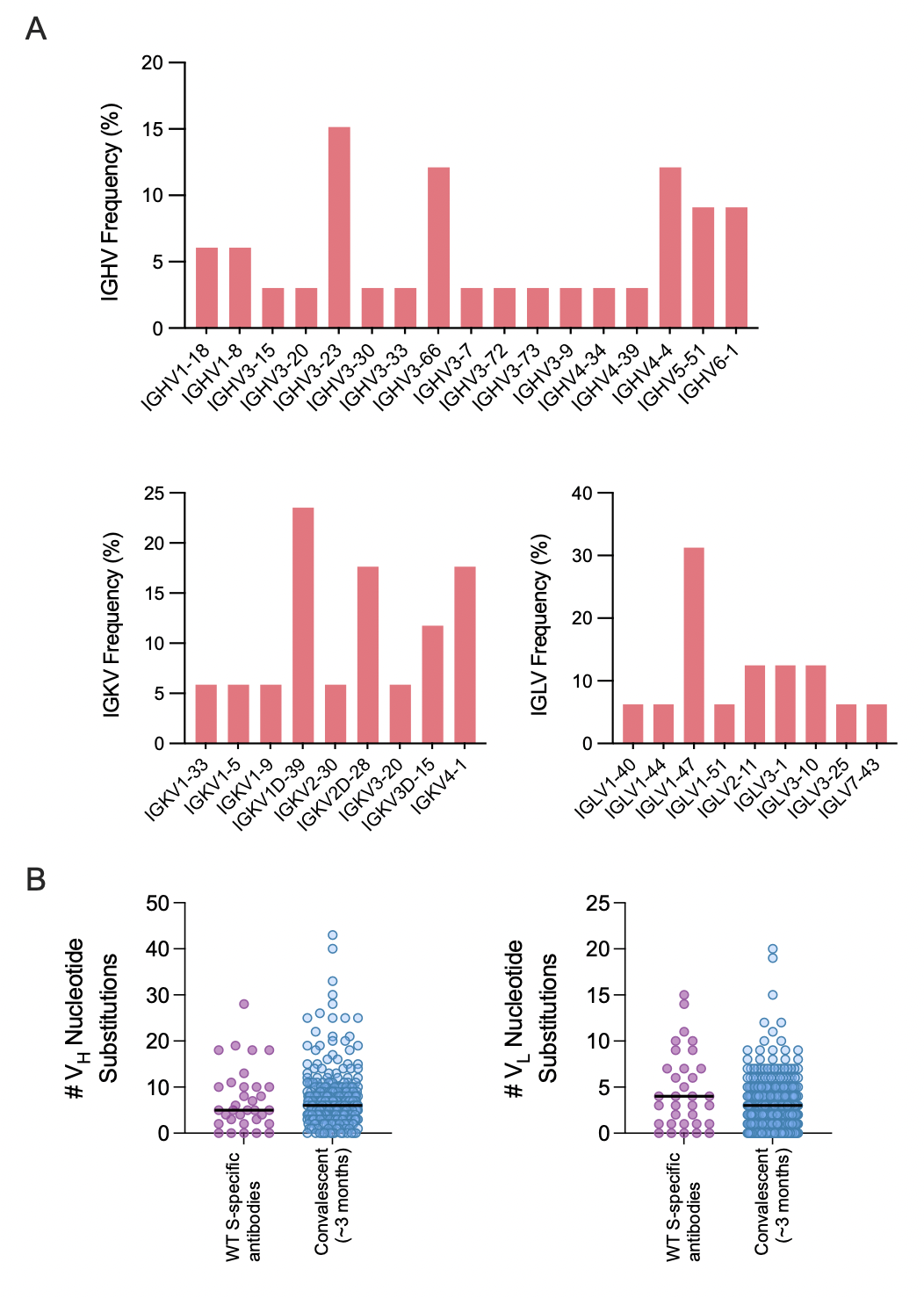

**Fig. S8.** Sequence features of WT S-specific antibodies. **(A)** IGHV, IGKV, and IGLV germline gene frequencies among WT-specific antibodies. **(B)** Number of somatic nucleotide mutations in the VH (left) and VL (right) genes in antibodies obtained from WT-specific B cells. SHM loads in S-specific antibodies obtained from SARS-CoV-2 convalescent individuals approximately 3 months post-infection are shown for comparison (*27*). Red bars indicate medians. IGHV, immunoglobulin heavy variable domain; IGKV, immunoglobulin kappa variable domain; IGLV, immunoglobulin lambda variable domain.

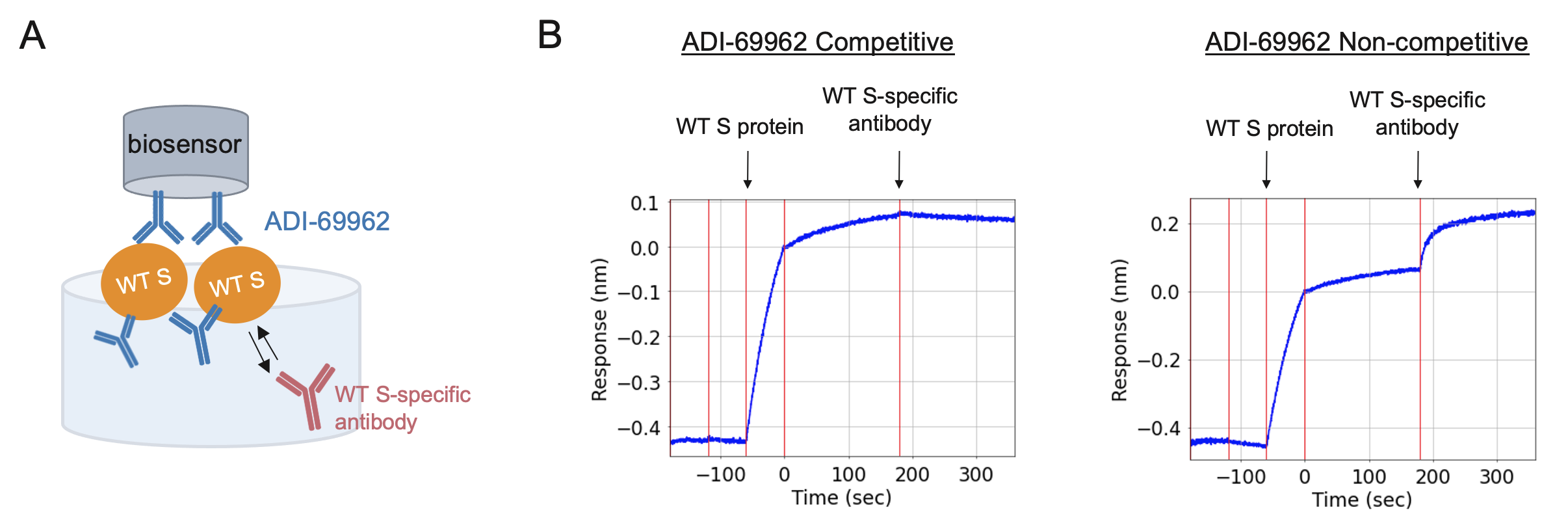

**Fig. S9.** ADI-69962 competitive binding assay. **(A)** Schematic of BLI assay set-up. Biosensors coated with the S2-directed antibody ADI-69962 were exposed to recombinant WT S. Remaining ADI-69962 epitopes present on WT S were blocked by soaking sensors in ADI-69962 before exposure to WT S-specific antibodies. **(B)** Representative BLI plots showing competitive (left) and non-competitive (right) binding profiles. Arrows above sensorgrams indicate exposure of ADI-69962-loaded biosensors to solutions containing WT S protein and WT S-specific antibodies.

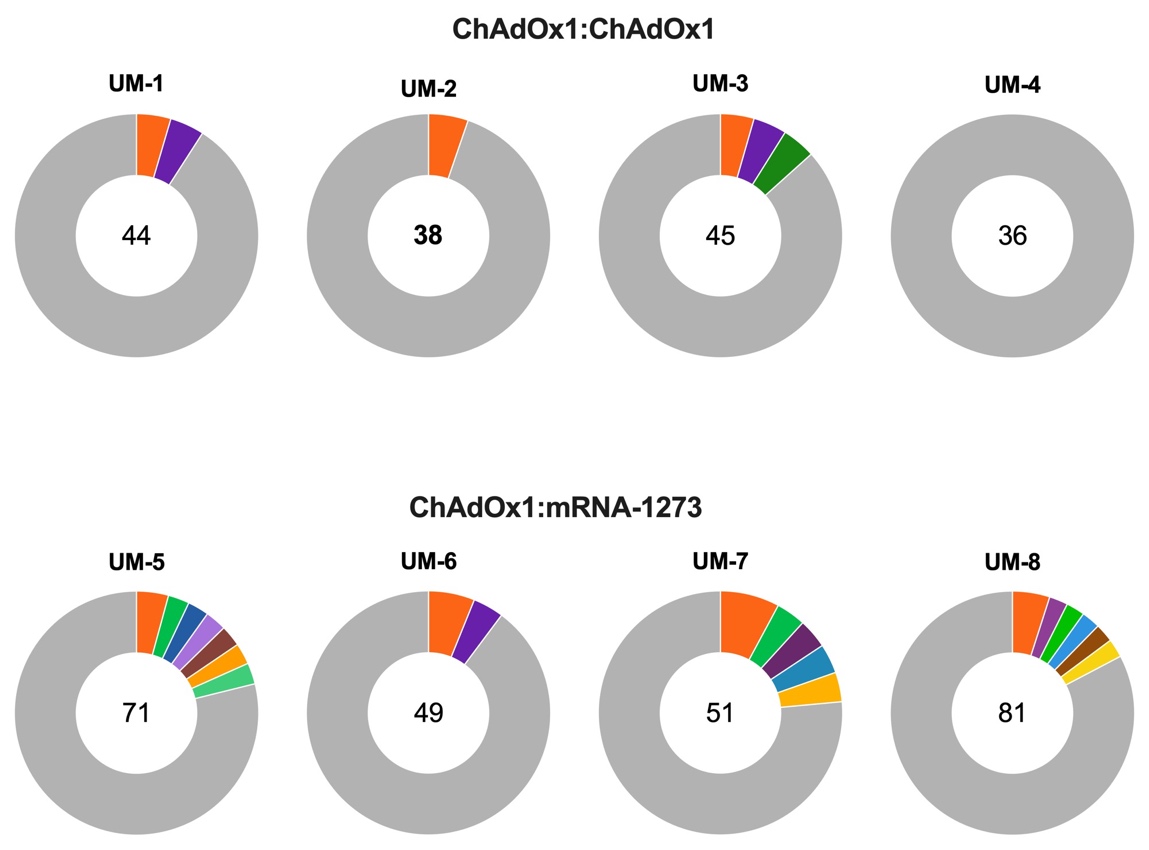

**Fig. S10.** Clonal lineage analysis of monoclonal antibodies isolated from ChAdOx1 (top) or mRNA-1273 (bottom) boosted donors. The size of the slice is proportional to the number of clones in that lineage, and number in the center of the pie indicates the total number of antibodies isolated from each donor. Unique clones are pooled and shown as a single grey segment. Clonal lineages were defined based on the following criteria: identical VH and VL germline gene, identical CDR H3 length, and ≥80% identity in CDR H3 amino acid sequence.

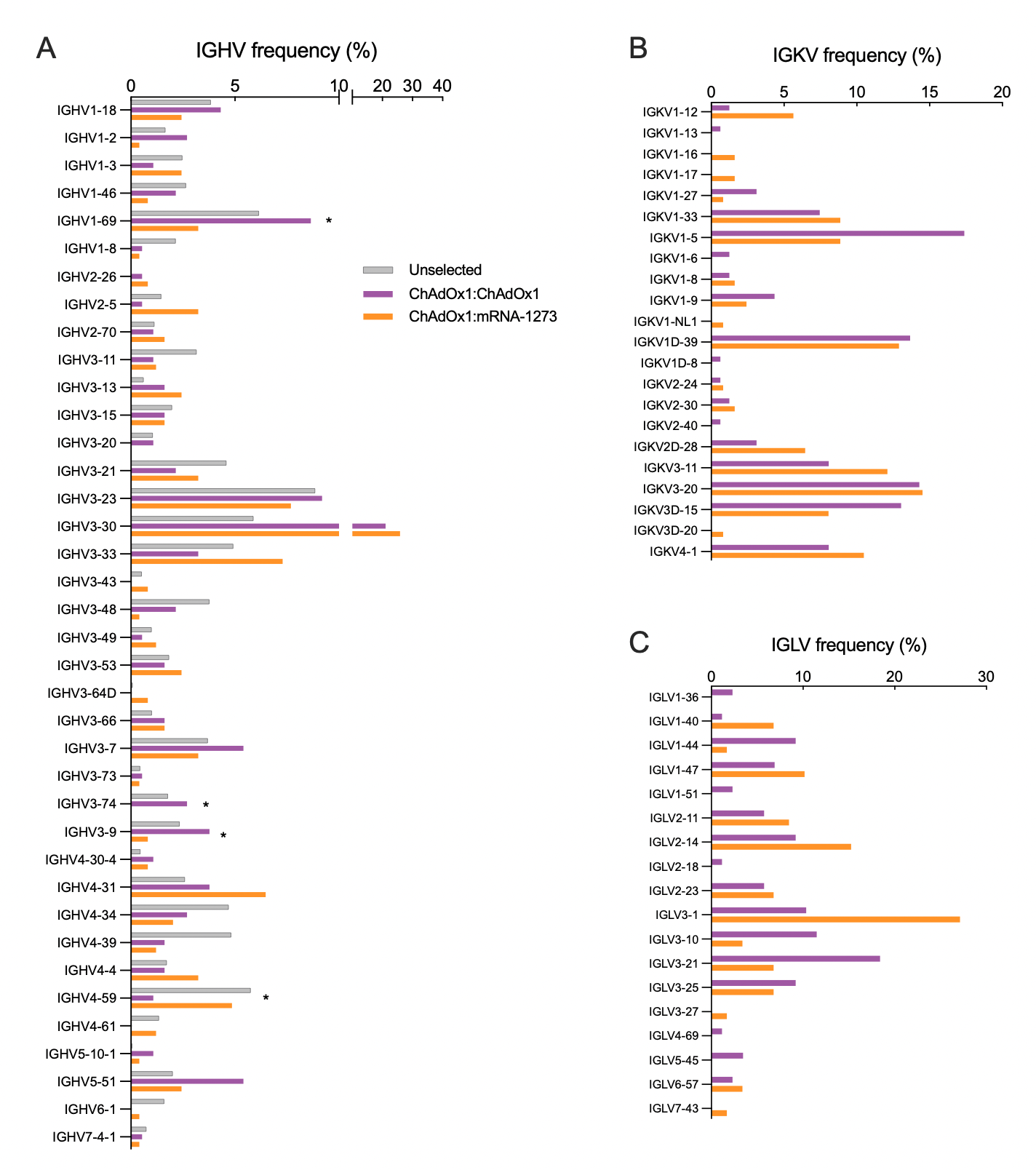

**Fig. S11.** Germline gene usage of S-2P-reactive antibodies isolated following homologous and heterologous immunization. **(A)** IGHV germline gene frequencies. Baseline (unselected) IGHV frequencies (grey) obtained by high-throughput sequencing of healthy donor B cells are included for reference (*29*). **(B-C)** Light chain germline gene frequencies for kappa (B) and lambda (C) chains. Statistical comparisons between donor cohorts were determined by Fisher's exact tests. *P<0.05. IGHV, immunoglobulin heavy variable domain; IGKV, immunoglobulin kappa variable domain; IGLV, immunoglobulin lambda variable domain.

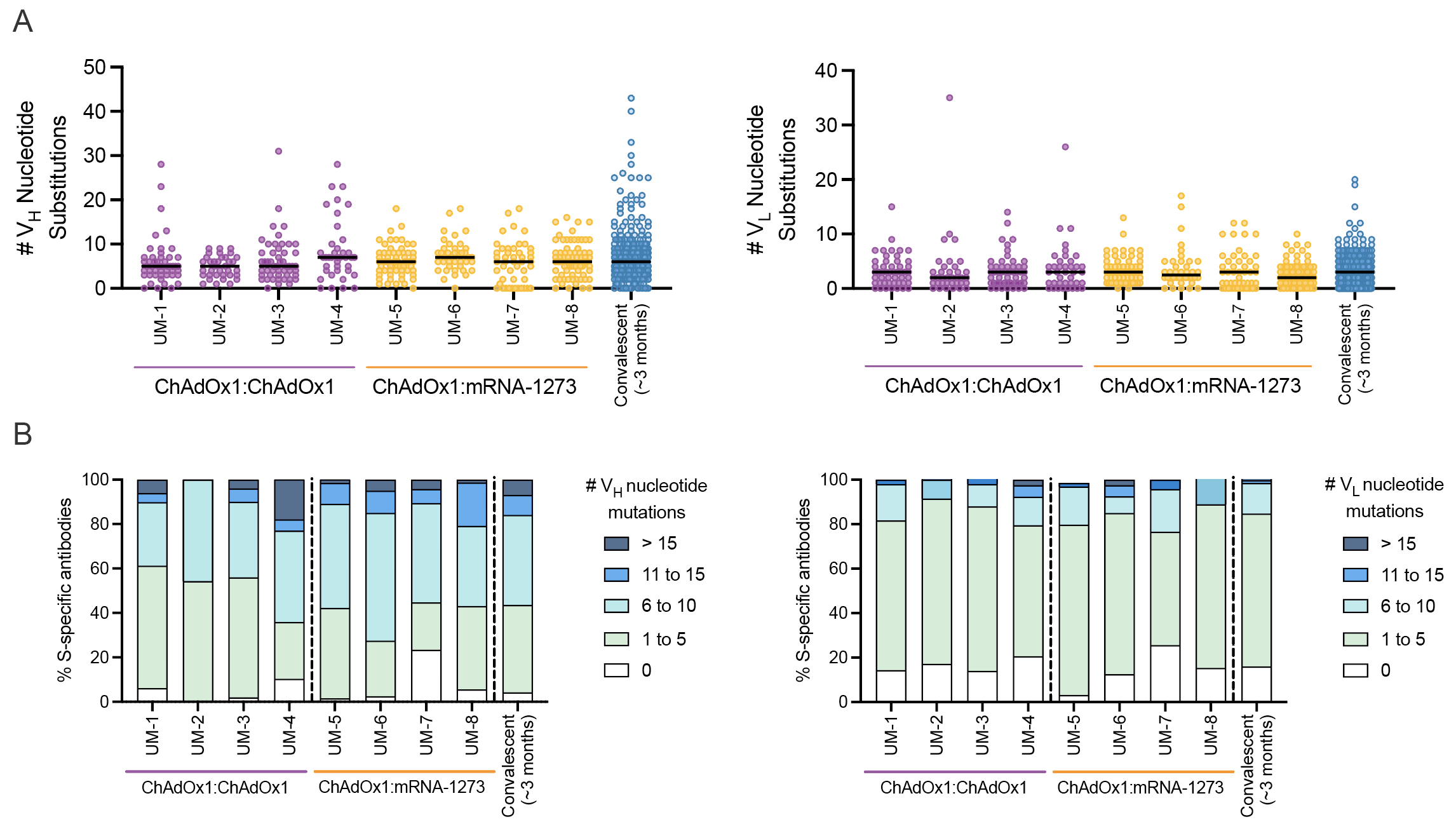

**Fig. S12.** Somatic hypermutation load in S-2P-reactive antibodies isolated from ChAdOx1:ChAdOx1 and ChAdOx1:mRNA-1273 vaccinated donors. **(A)** Number of nucleotide substitutions in VH (left) and VL (right) for antibodies isolated from each donor. Medians are shown as red bars. **(B)** Proportion of antibodies from each donor with the indicated number of nucleotide substitutions in the VH (left) and VL (right). SHM loads in antibodies isolated from SARS-CoV-2 convalescent individuals approximately 3 months post-infection are shown for comparison (*27*). VH, variable heavy chain; VL, variable light chain.

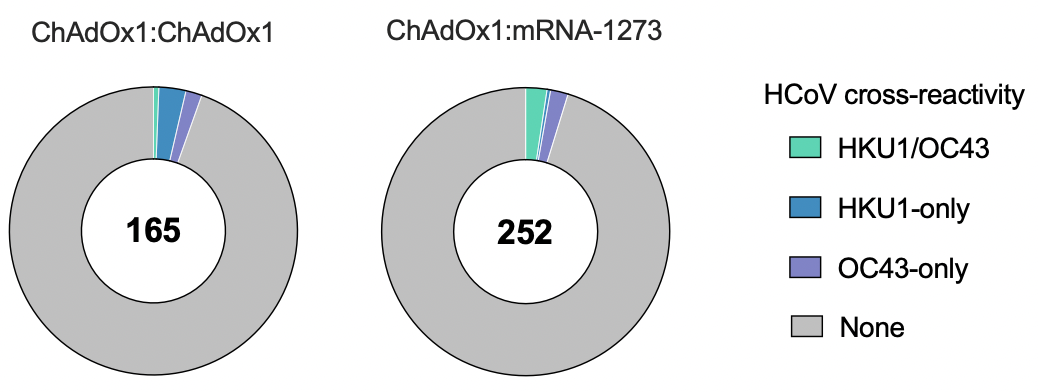

**Fig. S13.** Endemic β-CoV cross-reactivity of antibodies derived from ChAdOx1 and mRNA-1273 boosted donors. Colored slices indicate the proportion of antibodies reactive with HKU1 and/or OC43 S, as determined by BLI. The total number of antibodies analyzed is indicated in the center of the pie.

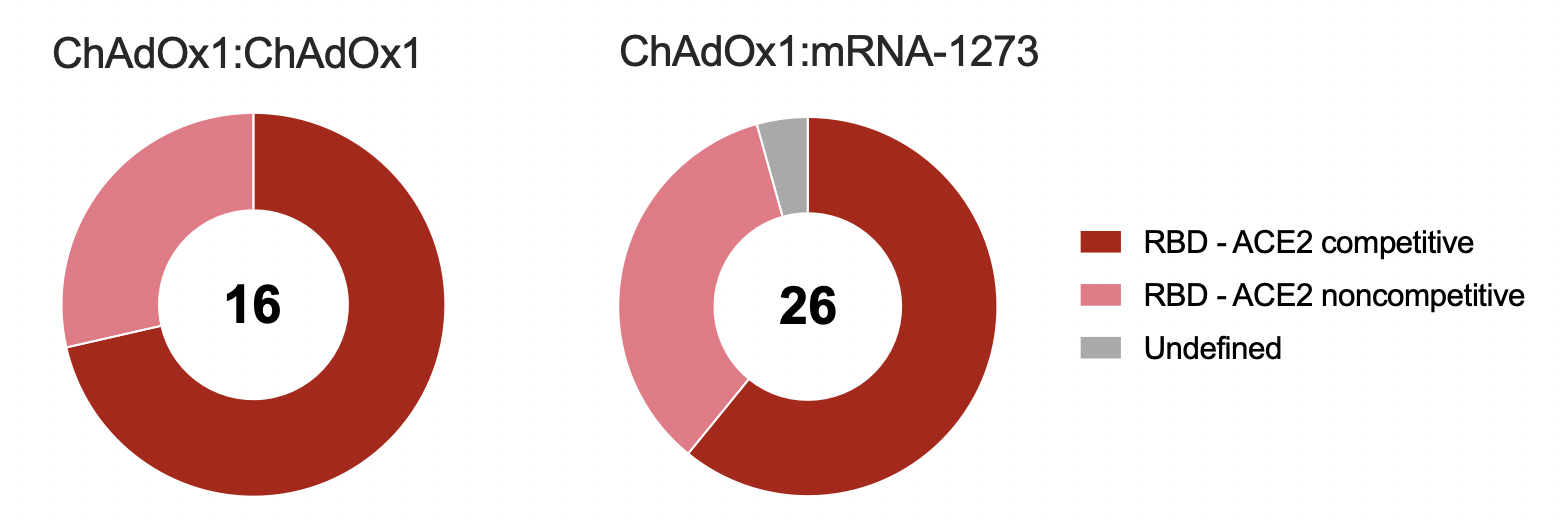

**Fig. S14.** Antigenic sites targeted by neutralizing antibodies derived from ChAdOx1 and mRNA-1273 boosted donors. Antibodies displaying >50% neutralization activity against MLV-SARS-CoV-2/Wuhan-1 at a concentration of 1 µg/ml are included in this analysis. The total number of neutralizing antibodies is indicated in the center of each pie.

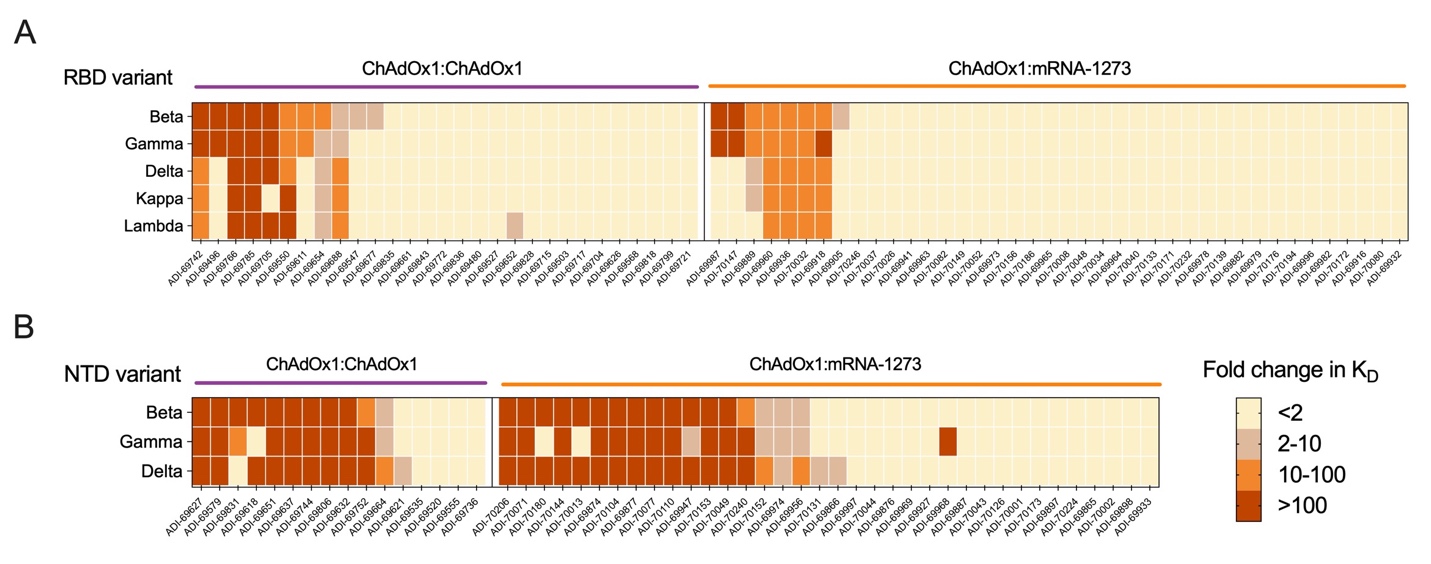

**Fig. S15.** Monoclonal antibody binding to variant RBD and NTDs. **(A-B)** Fold change in Fab binding affinity (K_D_) to variant RBDs (A) and NTDs (B) relative to the Wuhan-1 RBD and NTD, as determined by flow cytometry. K_D,_ equilibrium dissociation constant.

**Table S1.** Summary of donor characteristics.

|  | **ChAdOx1:ChAdOx1 (n=28)** | **ChAdOx1:mRNA-1273 (n=27)** |
| --- | --- | --- |
| **Age, median (range)** | 46.5 (28-62) | 42 (28-55) |
| **Sex no. (%)** |  |  |
| Male | 7 (25.0) | 8 (29.6) |
| Female | 21 (75.0) | 19 (70.4) |
| **BMI, median (range)** | 25.8 (19.3-36.0) | 22.9 (19.4-31) |
| **Comorbidities no. (%)** |  |  |
| Cardiovascular disease | 2 (7.1) | 0 |
| Diabetes mellitus type II | 1 (3.6) | 0 |
| Hypertension | 3 (10.7) | 0 |
| Asthma | 2 (7.1) | 3 (11.1) |
| Rheumatic disease | 0 | 1 (3.7) |
| Allergy | 6 (21.4) | 1 (3.7) |
| **Days between doses median (range)** | 70 (60-80) | 84 (59-87) |

**Table S2.** Donors selected for monoclonal antibody isolation.

| Subject | Age range | Sex | Immunization regimen (prime:boost) | Days between doses |
| --- | --- | --- | --- | --- |
| UM-1 | 51-55 | Female | ChAdOx1:ChAdOx1 | 60 |
| UM-2 | 31-35 | Male | ChAdOx1:ChAdOx1 | 76 |
| UM-3 | 56-60 | Female | ChAdOx1:ChAdOx1 | 60 |
| UM-4 | 36-40 | Female | ChAdOx1:ChAdOx1 | 72 |
| UM-5 | 51-55 | Female | ChAdOx1:mRNA-1273 | 82 |
| UM-6 | 41-45 | Female | ChAdOx1:mRNA-1273 | 83 |
| UM-7 | 41-45 | Male | ChAdOx1:mRNA-1273 | 86 |
| UM-8 | 41-45 | Female | ChAdOx1:mRNA-1273 | 86 |
